# Cost-effectiveness and health impact of gender-neutral and single-dose HPV vaccination in Hong Kong

**DOI:** 10.1101/2025.10.20.25338351

**Authors:** Horace Cheuk Wai Choi, Kathy Leung, Mark Jit, Joseph T Wu

**Affiliations:** WHO Collaborating Centre for Infectious Disease Epidemiology and Control, School of Public Health, Li Ka Shing Faculty of Medicine, The University of Hong Kong, Hong Kong SAR, China; Laboratory of Data Discovery for Health Limited (D24H), Hong Kong SAR, China; The Hong Kong Jockey Club Global Health Institute, Hong Kong SAR, China; The University of Hong Kong – Shenzhen Hospital, Shenzhen, China; Department of Global and Environmental Health, School of Global Public Health, New York University, New York, United States of America; Department of Infectious Disease Epidemiology & Dynamics, London School of Hygiene & Tropical Medicine, London, United Kingdom; Saw Swee Hock School of Public Health, National University of Singapore, Singapore

**Keywords:** Gender-neutral vaccination, cost-effectiveness analysis, nonavalent HPV vaccines, population-based HPV vaccination

## Abstract

**Background:** Since 2019, Hong Kong has implemented a routine nonavalent human papillomavirus (HPV) vaccination program for schoolgirls aged 10-12 years with two-dose uptake of over 85%. However, the impacts of gender-neutral vaccination (GNV) with a single-dose schedule have not been studied.

**Objective:** To evaluate the cost-effectiveness of expanding the two-dose female-only vaccination (2dFOV) to GNV and reducing the schedule to one dose.

**Methods:** We modeled the impacts of HPV vaccination on the burden of HPV-related cancers in both genders at various vaccine uptake among schoolboys. We estimated the changes in the associated costs and health benefits across the lifetime of all cohorts of both genders over a time horizon of 100 years with a 3% annual discount rate. We calculated the incremental cost-effectiveness ratio (ICER) of expanding 2dFOV to GNV, using a two-dose or one-dose schedule compared to a threshold of one gross domestic product per capita (US$48,757). Sensitivity analyses were performed to assess the uncertainty of the findings.

**Results:** Assuming base case vaccination cost and 85% uptake for both genders, two-dose GNV (2F2M) has an ICER of US$109,375 (90% prediction interval: (US$63,824, US$264,980)) compared to 2dFOV and is not cost-effective. Compared to 2dFOV under the same assumptions, giving one dose to both genders (1F1M) always results in QALY gains if one-dose schedule provides 30-year protection; this strategy is cost-effective (and may be cost-saving). If the one-dose schedule gives only 20-year protection, 1F1M incurs QALY gains in 91% and 67% of simulations if boys’ uptake is 85% and 50%, respectively. Compared to 2dFOV, if 85% of boys are vaccinated and one-dose schedule provides at least 20 years of protection, adding one dose for boys (2F1M) is cost-effective in 47% of simulations at the base case vaccination cost.

**Conclusions:** 1F1M is more effective than 2dFOV if it protects for at least 20-30 years with boys’ uptake of 50% or above. The findings highlight the potential of implementing GNV with a single-dose schedule for better resource allocation and optimizing the impacts of the vaccination program.

## Introduction

Persistent high-risk oncogenic human papillomavirus (HPV) infection is a necessary cause of cervical cancer and can also lead to non-cervical cancers including anal, oropharyngeal, vaginal/vulvar, and penile cancers.^[1,2]^ Hence, both females and males can directly benefit from receiving HPV vaccines for preventing HPV infection and subsequent development of HPV-related cancers.^[3,4]^

In its position paper updated in 2022, the World Health Organization (WHO) reviewed the use of HPV vaccination with a primary focus on accelerating the elimination of cervical cancer (i.e., bringing incidence below 4 per 100,000 women-years).^[5,6]^ The WHO recommended a two-dose or single-dose schedule for girls aged 9-14 years who are the primary target population of HPV vaccination. Older females and boys are the secondary target populations if such vaccinations are feasible and affordable. As of May 2025, over 140 countries/regions have initiated population-based HPV vaccination programs for female adolescents, and over 70 countries have implemented the single-dose schedule.^[7]^ Moreover, more than 80 countries have extended female-only vaccination (FOV) to gender-neutral vaccination (GNV) in their population-based HPV vaccination programs for adolescents. Some of these countries, such as Australia and the United Kingdom, have moved to a single-dose schedule for both genders since 2023.^[8,9]^

Since 2019, Hong Kong has implemented a population-based HPV vaccination program for schoolgirls in primary five and six (aged 10-12 years).^[10]^ The program achieved uptake of over 85% in 2023 among targeted schoolgirls for the two-dose nonavalent HPV (9vHPV) vaccine.^[11]^ In response to recent global developments in HPV vaccination, we evaluated the cost-effectiveness of potentially updating the current HPV vaccination program in Hong Kong by expanding FOV to GNV, and of adopting a single-dose schedule.

## Methods

### Models

To estimate the change in HPV infection prevalence and the impacts associated with large-scale GNV, we adapted our previous model that was used to assess the cost-effectiveness of schoolgirl 9vHPV vaccination in Hong Kong.^[12]^ The model included an HPV transmission model with four groups of HPV types (namely, HPV-16, HPV-18, HPV-31/33/45/52/58, and other non-vaccine high-risk HPV types), and an individual-based model to account for the effect of cervical screening. The model was calibrated to the local age- and type-specific genital HPV prevalence among females and age-specific cervical cancer incidence. Supplementary 1 (S1.1) provides more details of the model.

We expanded this model by considering oropharyngeal cancers (OPCs), anal cancers, penile cancers, and vaginal/vulvar cancers when evaluating the health impacts of HPV vaccination. Data on the incidence of these cancers (stratified by age and sex) were obtained from the Hong Kong Cancer Registry (HKCaR).^[13]^ A local study reported that high-risk HPV infection was present in 42% (95% confidence interval [CI]: 35% to 49%) of 179 OPC cases diagnosed in 2016-2020.^[14]^ However, more detailed local data on high-risk HPV prevalence by age and HPV attributable fraction (i.e., the proportion of cancer cases that are associated with high-risk HPV infection) at non-cervical sites are limited. In the absence of such data, to estimate the impacts of HPV vaccination on non-cervical HPV-related cancers, we assumed that for each gender, age-specific incidence of each of these cancers is statistically associated with age-specific genital HPV infection incidence via a three-parameter time-lagged link function.^[15,16]^ We fitted the model to empirical cancer incidence from HKCaR and the reported relative prevalence of high-risk HPV types among HPV-positive cancer cases in the literature.^[2,13,14]^ Using the fitted model, we projected the changes in non-cervical HPV-related cancer incidence following different strategies of HPV vaccination. See Supplementary 1 (S1.2.-S1.3.) for more details on model calibration and estimation of non-cervical HPV-related cancer incidence.

### HPV Vaccination

We used the current two-dose FOV program (2dFOV) in Hong Kong as the comparator.^[10]^ We then considered GNV scenarios that extend 2dFOV to vaccinating schoolboys in primary five/six using 9vHPV vaccines. We assumed that 9vHPV vaccines provide the same HPV type-specific efficacy of 95% in preventing vaccine-targeted HPV infection (HPV-16/18/31/33/45/52/58) in both genders,^[12,17,18]^ but no efficacy against non-vaccine HPV types.

We assumed that vaccine uptake of primary five/six schoolgirls would remain at 85% in all scenarios based on the most recent coverage data.^[11]^ We considered three scenarios of vaccine uptake among schoolboys of 85%, 50%, and 25% based on the observed uptake among schoolgirls and local surveys of acceptability of male adolescent HPV vaccination.^[19,20]^ We evaluated three GNV strategies: (a) two doses for both schoolgirls and schoolboys (2F2M), (b) one dose for both genders (1F1M), and (c) two doses for schoolgirls and one dose for schoolboys (2F1M). To reflect the most pessimistic scenarios about the effectiveness of one-dose vaccination, we assumed that the two-dose schedule provided lifelong protection while the one-dose schedule provided only 20 or 30 years of shorter protection.^[21,22]^

### Cost-effectiveness analysis (CEA)

We conducted the CEA using a societal perspective. We estimated the costs of cancer treatment based on the corresponding charges for private care in the Government’s Gazette by the Hospital Authority.^[12,23]^ We included costs of cervical screening for females, assuming 70% screening attendance among women aged 25-64 years, with HPV testing and cytology as the primary modality for women aged 30-64 and 25-29 years, respectively.^[24,25]^ In the base case, we set the cost of vaccination at US$218 per dose (including vaccine cost and administration expenses).^[26]^ We quantified health outcomes using quality-adjusted life years (QALYs).^[27]^ We estimated the changes in the associated costs and QALYs across the lifetime of all cohorts of both genders over a time horizon of 100 years with a 3% annual discount rate.^[12]^

We assessed cost-effectiveness by estimating the incremental cost-effectiveness ratio (ICER) of GNV, defined as the incremental cost divided by the incremental QALYs gained from expanding FOV to GNV. We considered one gross domestic product per capita (GDPpc; US$48,757) per QALY gained as the willingness to pay (WTP) threshold, as done in previous health technology assessments of a similar nature in Hong Kong.^[12,28,29]^ This WTP threshold paralleled the estimated cost-effectiveness thresholds among high-income countries that have a similar life expectancy and GDPpc to Hong Kong.^[30]^ We also evaluated the threshold vaccination cost (TVC) for expanding FOV to GNV, defined as the cost per dose of vaccination at which GNV and FOV would be equally cost-effective at the WTP threshold of one GDPpc. We assumed that the vaccination cost was the same for both genders.

We used probabilistic sensitivity analysis to account for parametric uncertainty in the natural history of cervical cancer and HPV transmission dynamics, the statistical linkages between genital HPV infections and non-cervical HPV-related cancers, as well as costs and health utilities. Outcomes in terms of ICERs and TVCs are summarized using medians and 90% prediction intervals (PIs; 5^th^ to 95^th^ percentiles). To avoid overestimating the health benefits of GNV, we assumed in the base case that all HPV-related anal cancers in males are equally avertable by FOV via indirect protection. In the sensitivity analysis, we considered an alternative scenario in which only 60% of HPV-related anal cancers in males were avertable by FOV, based on the proportion of male anal cancer patients who were men who have sex with men (MSM) in the United States.^[31]^ Furthermore, we conducted a sensitivity analysis to take into account the impact of HPV vaccination on genital warts, assuming that 90% of genital warts were attributable to HPV types 6/11 which are preventable by 9vHPV vaccines.^[32,33]^ See S2 in Supplementary 1 for details of the CEA.

In view of WHO’s cervical cancer elimination initiative, we estimated the time it takes for the age-standardized incidence rate (ASIR) for cervical cancer to fall below 4 per 100,000 women-years following the implementation of the population-based HPV vaccination program.^[6,13]^

Model simulations were performed using Matlab, C++, and R. We followed the Consolidated Health Economic Evaluation Reporting Standards (CHEERS) 2022 statement and HPV-FRAME checklist for reporting health economic and HPV-related cancer control evaluations, respectively.^[34,35]^

## Results

### CEA

#### 2F2M

Compared to 2dFOV, 2F2M was not cost-effective. Indeed, ICER increased with higher vaccine uptake among boys (Figure 1). In the base case scenario, assuming vaccine uptake among boys mirrors the current uptake among girls (85%), the ICER of 2F2M was US$109,375 (US$63,824, US$264,980) and the TVC was US$107 (US$46, US$174) (eTable 3). At this uptake, 2F2M was not cost-effective in 99% of simulations at the base case vaccination cost. At 85% uptake, 2F2M remained not cost-effective in 97% of simulations at the base case vaccination cost even if only 60% of HPV-related anal cancers in males were avertable by FOV and the burden of genital warts was included in the CEA.

**Figure 1.**
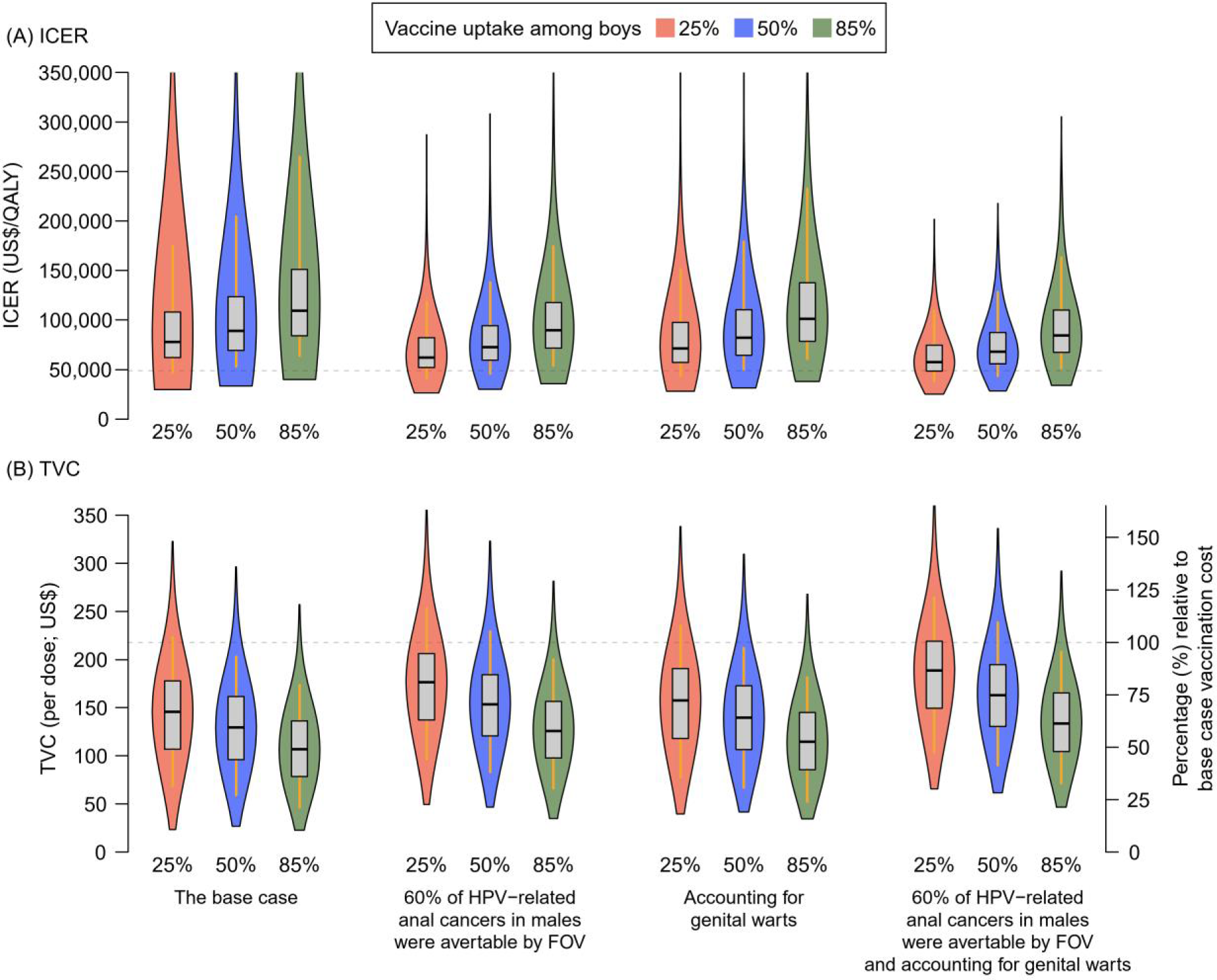
Incremental cost-effectiveness ratios (ICERs; A) and threshold vaccination costs (TVCs; B) for 2F2M vs 2dFOV. Abbreviations: FOV, female-only vaccination; GNV, gender-neutral vaccination; 2dFOV, two-dose schedule for female-only vaccination; 2F2M, two-dose schedule for both schoolgirls and schoolboys. ^a^ The two-dose schedule was assumed to provide lifelong protection to vaccinees. ^b^ Vaccine uptake among girls was 85% in all GNV and FOV strategies. ^c^ For panel (A), the willingness to pay (WTP) threshold was US$48,757 per QALY gain. For panel (B), the base case vaccination cost (vaccine cost plus administration expenses) was US$218 per dose. The WTP threshold and base case vaccination cost were presented by grey dashed lines on respective panels. ^d^ The violin plots present the smoothed kernel density of the values of ICERs/TVCs. The box plots present the medians (the horizontal lines), the 25^th^/75^th^ percentiles (the boxes), and the 5^th^/95^th^ percentiles (the whiskers).

#### 1F1M

The impact of 1F1M was sensitive to vaccine uptake in boys and the protection duration of the one-dose schedule. If the one-dose schedule provided 20 years of protection, 1F1M may incur QALY losses compared to 2dFOV. In the base case, 1F1M generated fewer QALYs than 2dFOV in 75% and 9% of simulations if vaccine uptake among boys was 25% and 85%, respectively (Figure 2A(i)). However, if the one-dose schedule provided 30-year protection, 1F1M generated more QALYs in most simulations regardless of vaccine uptake in boys (Figure 2B). In the base case, if schoolboys’ uptake was 85%, 1F1M generated QALY gains in all simulations and was either cost-effective (31%) or cost-saving (69%) (Figure 2B(i)).

**Figure 2.**
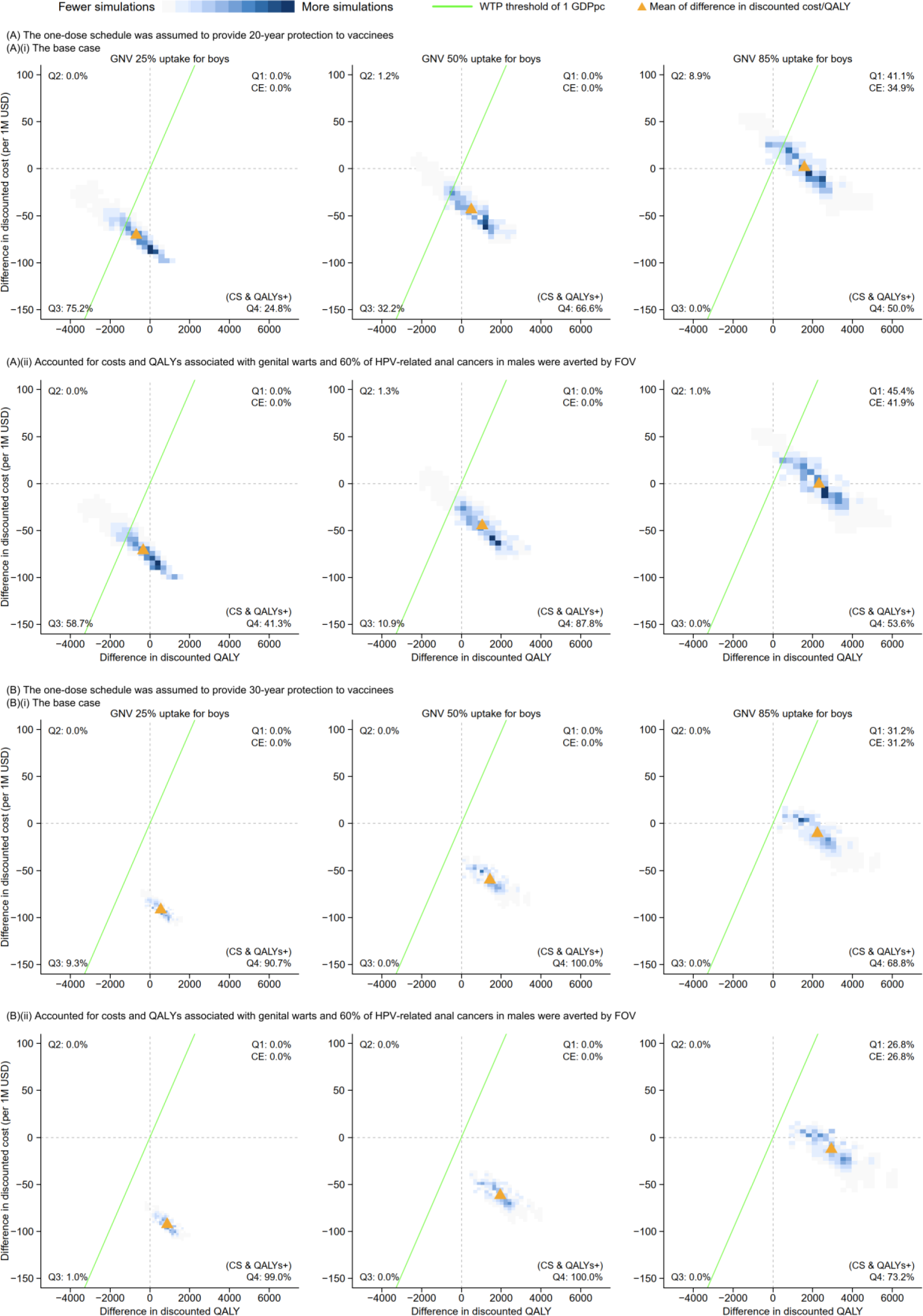
Cost-effectiveness planes for 1F1M vs 2dFOV if the one-dose schedule was assumed to provide (A) 20-year and (B) 30-year protection. Abbreviations: CE, cost-effective; CS: cost-saving; QALY, quality-adjusted life year; Q1 to Q4, quadrant 1 to quadrant 4 of the cost-effectiveness plane; 1F1M, one-dose schedule for both schoolgirls and schoolboys; 2dFOV, two-dose schedule for female-only vaccination. ^a^ The two-dose schedule was assumed to provide lifelong protection. The duration of vaccine-induced protection of the one-dose schedule was assumed to be 20 years in panel (A) and 30 years in panel (B). ^b^ Vaccine uptake among girls was 85% in all GNV and FOV schedules. ^c^ The green lines present the willingness to pay (WTP) threshold at US$48,757/QALY. The orange triangles denote the mean of the differences in discounted cost and QALY. ^d^ On the density heatmaps (2-D histograms), grey/lighter blue represents fewer simulations and darker blue represents more simulations. ^e^ In each subplot, the percentage of simulations in each quadrant (i.e., Q1-Q4) is presented. Quadrant Q1 (North-east, higher costs and more QALYs) includes the percentage of simulations that were cost-effective at the WTP threshold. All percentages are based on the total number of simulations.

#### 2F1M

Although dose schedules are typically agnostic to gender in routine immunization programs, we explored the potential impact of adopting 2F1M as an alternative to 2dFOV or 1F1M. The ICER of 2F1M compared to 2dFOV increased as vaccine uptake among boys increased (Figure 3/eFigure 6). The cost-effectiveness outcomes of 2F1M were not sensitive to the duration of protection provided by the one-dose schedule as long as the protection was between 20 and 30 years (eTable 4-5). 2F1M was a cost-effective alternative to 2dFOV if the vaccination cost could be moderately reduced. In the base case scenario with 85% uptake among boys, 2F1M was cost-effective in 47% of simulations at the base case vaccination cost and became cost-effective in 80% of simulations if the vaccination cost was reduced by 33% (eTable 4-5). In contrast, compared to 1F1M, 2F1M was not cost-effective in most simulations across all considered scenarios (eFigure 7-8/eTable 6-7).

**Figure 3.**
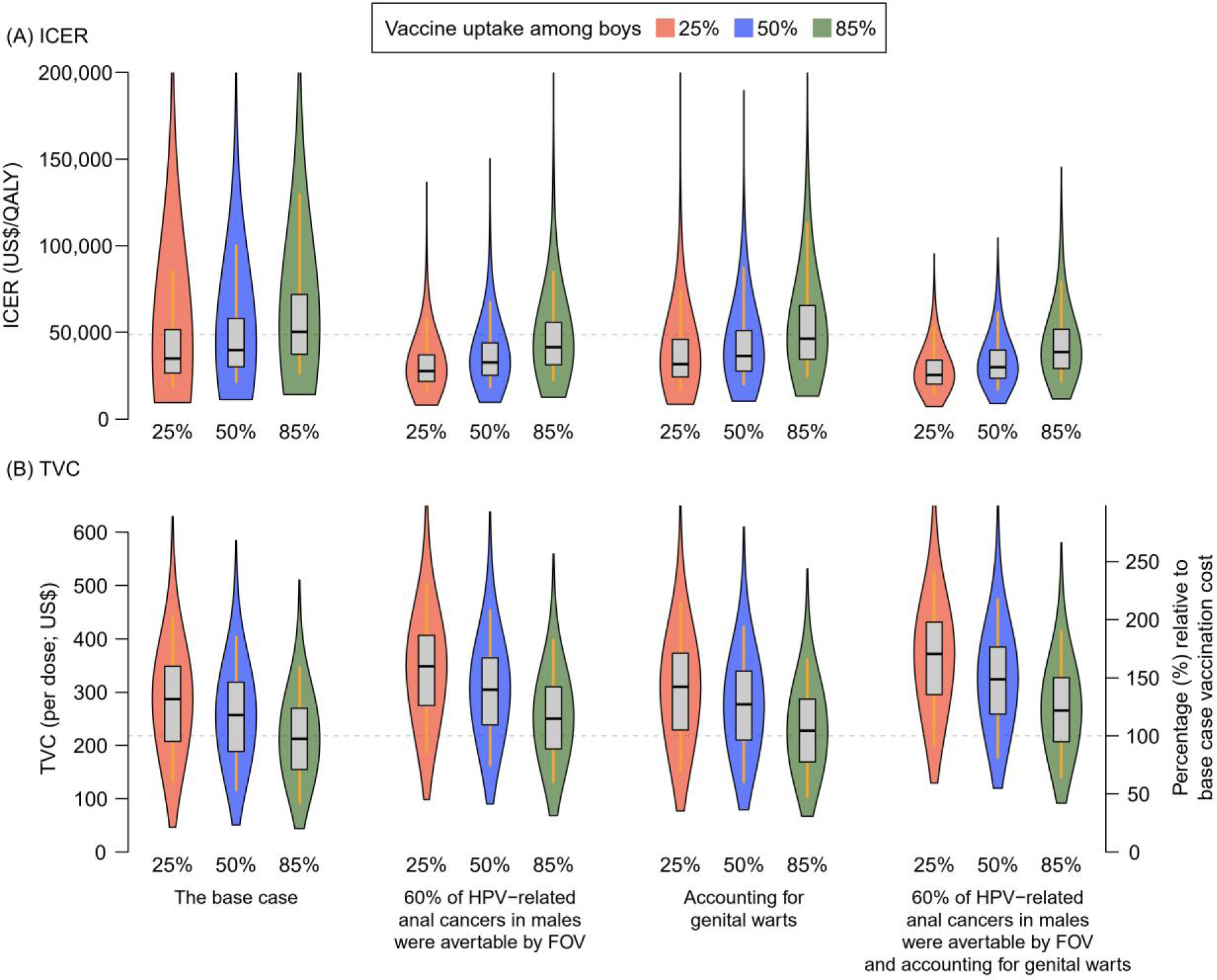
Incremental cost-effectiveness ratios (ICERs; A) and threshold vaccination costs (TVCs; B) for 2F1M vs 2dFOV if the one-dose schedule provided 20-year protection to vaccinees. Abbreviations: FOV, female-only vaccination; GNV, gender-neutral vaccination; 2F1M, two-dose vaccination schedule for schoolgirls and one-dose for schoolboys; 2dFOV, two-dose schedule for female-only vaccination. ^a^ The two-dose and one-dose schedules were assumed to provide lifelong and 20-year protection to vaccinees, respectively. ^b^ Vaccine uptake among girls was 85% in all GNV and FOV strategies. ^c^ For panel (A), the willingness to pay (WTP) threshold was US$48,757 per QALY gain. For panel (B), the base case vaccination cost (vaccine cost plus administration expenses) was US$218 per dose. The WTP threshold and base case vaccination cost were presented by grey dashed lines on respective panels. ^d^ The violin plots present the smoothed kernel density of the values of ICERs/TVCs. The box plots present the medians (the horizontal lines), the 25^th^/75^th^ percentiles (the boxes), and the 5^th^/95^th^ percentiles (the whiskers).

### Projected timeline for the elimination of cervical cancer

We estimated that under the status quo 2dFOV (85% vaccine uptake in schoolgirls) and cervical screening program (70% screening uptake among women aged 25-64 years), Hong Kong would eliminate cervical cancer in 2067 (90% PI: (2064, 2070)) (Figure 4). 2F2M with 85% vaccine uptake among boys would eliminate cervical cancer 2 years sooner in 2065 (2062, 2068). However, if the 2dFOV program was complemented with an increase in screening uptake from 70% to 85% (as reported in a recent local survey),^[25]^ the elimination target could be reached in 2058 (2056, 2060), i.e., 9 (8, 10) years sooner. These findings suggest that increasing screening uptake might have a stronger marginal impact on the timeline of cervical cancer elimination in Hong Kong than expanding FOV to GNV.

**Figure 4.**
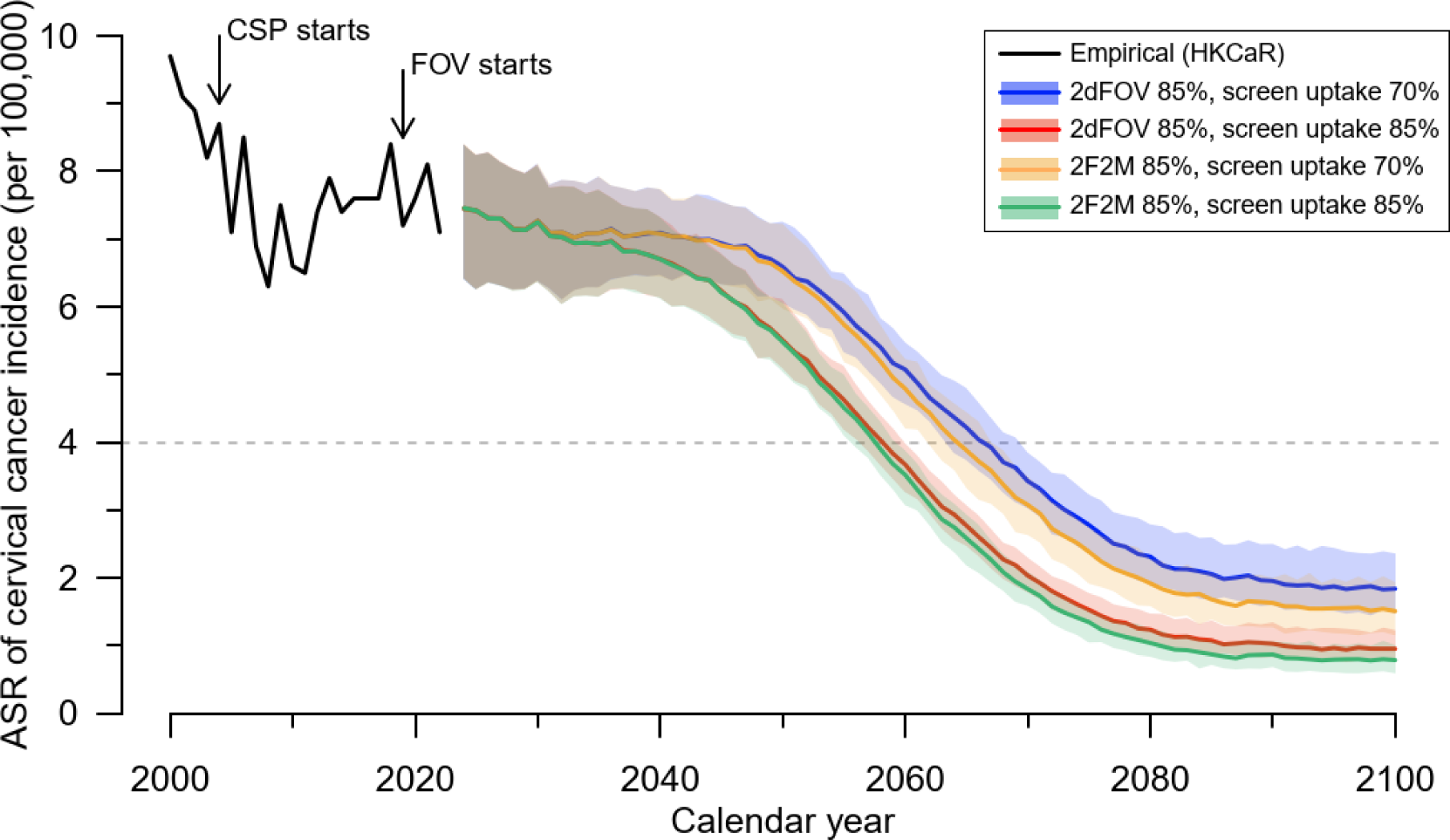
Estimated age-standardized rate of cervical cancer incidence across time horizon. Abbreviations: ASR, age-standardized rate; CSP, Cervical Screening Programme; HKCaR, Hong Kong Cancer Registry; 2dFOV, female-only HPV vaccination with a two-dose schedule for schoolgirls; 2F2M, gender-neutral HPV vaccination with a two-dose schedule for both schoolgirls and schoolboys. ^a^ Lifelong vaccine protection was assumed for a two-dose schedule of 2dFOV and 2F2M. ^b^ The age-standardized incidence rate (ASIR) of cervical cancer was calculated based on Segi’s 1960 world standard population.^[13]^ ^c^ Empirical ASIR (the black line) from the Hong Kong Cancer Registry was available up to 2022.^[13]^ ^d^ The blue and red lines (shaded regions) present the medians (90% prediction intervals [PIs]; 5^th^ to 95^th^ percentiles) of the estimated ASIR for 2dFOV with vaccine uptake of 85% among girls when screening rates were 70% and 85%, respectively. The orange and green lines (shaded regions) present the corresponding medians (90% PIs) for 2F2M with vaccine uptake of 85% among girls and boys when screening uptakes were 70% and 85%, respectively. The medians and 90% PIs were obtained based on probabilistic sensitivity analysis. ^e^ The grey dashed line presents the reference of cervical cancer elimination for ASIR at 4 per 100,000 women-years.

S3 of Supplementary 1 presents more findings on comparing GNV to 2dFOV. Considering the distribution of the difference in costs and QALYs gained associated with treatments of HPV-related cancers and genital warts, the greatest proportion of health impacts due to GNV are attributable to cervical cancer prevention, followed by male HPV-related cancer prevention (OPC and anal cancer) and genital warts (eFigure 9). If the vaccination schedule was changed from 2dFOV to 1F1M with 20- or 30-year protection from a one-dose schedule, a rebound in the incidence of vaccine-targeted HPV types is seen in older ages when vaccine-induced protection wanes (eFigure 13-14). The rebound of incidence occurs in later ages among males, and in scenarios of higher uptake among boys.

## Discussion

In this study, we evaluated the cost-effectiveness of expanding the current routine adolescent 9vHPV vaccination program in Hong Kong from FOV to GNV, accounting for the benefits of GNV in reducing cervical cancer incidence in women (due to herd effects) and non-cervical HPV-related cancer incidence in both genders.

At the current vaccine uptake (85%) of the FOV program,^[11]^ our study suggested that 2F2M would not be a cost-effective alternative to 2dFOV at the base case vaccination cost (US$218 per dose) under a WTP threshold of one GDPpc. In the base case scenario, the TVC was 51% (median) lower than the base case vaccination cost. This finding aligned with the results reported in other European and Asian countries, such as Spain, the United Kingdom, Japan, and Singapore, which found that 2F2M with 9vHPV vaccines was not cost-effective compared to 2dFOV.^[36–39]^ Specifically, the United Kingdom and Singaporean studies indicated that the vaccination cost needed to be reduced by 51% and 67%, respectively, for 2F2M using 9vHPV vaccines to be cost-effective.^[37,39]^

A recent manufacturer-sponsored local study assessed the cost-effectiveness of 2F2M with 9vHPV vaccines using the manufacturer’s model re-calibrated to the overall crude incidence rate of HPV-related cancers in Hong Kong.^[28]^ Assuming 80% vaccine uptake and accounting for genital warts, the study reported that 2F2M was cost-effective compared to 2dFOV with an ICER of US$40,500/QALY from a healthcare payer perspective. Compared to our study, their study assumed lower cervical screening coverage (13%), thereby making the health impact of HPV vaccination on reducing cervical cancer more pronounced, and lower vaccination cost (US$178 per dose). If we adopted their assumptions regarding vaccination and screening, our model estimated an ICER of US$41,100/QALY (median) for 2F2M vs 2dFOV.

Because 2F2M was unlikely to be cost-effective compared to 2dFOV, we considered the single-dose schedule, in line with WHO’s updated recommendation in 2022.^[5]^ Compared to 2dFOV with lifelong protection, a one-dose schedule with shorter protection duration may result in a rebound of incidence of vaccine-targeted HPV types, by delaying HPV infection from the age of peak sexual activity to older ages. Such a delay still reduces the risk of HPV-related cancers.^[22]^ The genital prevalence of high-risk HPV types among females in Hong Kong showed a major peak (approximately 10%) at ages 19-30 years and a lower peak (approximately 5%) with a steady decrease at ages 36-55 years.^[40,41]^ For a vaccination program targeting adolescents aged 10-12 years old, a one-dose schedule giving 20-year protection would cover the major peak of HPV infection. A longer protection duration of 30 years could also cover half of the age range of the lower peak. These findings highlight the influence of protection duration (20-year or 30-year) on the impact and cost-effectiveness of a one-dose schedule, even though moving from 2dFOV to 1F1M is likely to be cost-effective under either scenario.

In contrast to 1F1M, 2F1M was less sensitive to the protection duration of the one-dose schedule, as its cost-effectiveness compared to 2dFOV was similar under the assumptions of 20-year and 30-year protection. If a gender-specific vaccine schedule is deemed acceptable, 2F1M would be more cost-effective than 2dFOV although less cost-effective than 1F1M. Also, vaccine prices negotiated through tender-based procurement are usually lower than list prices,^[42]^ suggesting the potential to reach a moderate reduction in vaccination cost that enhances the cost-effectiveness of 2F1M.

Moreover, since 2021, two new HPV vaccines have gained WHO’s prequalification (Cecolin and Walrinvax).^[43]^ These vaccines have similar efficacy (using serological endpoints) to first-generation HPV vaccines,^[44,45]^ but are being sold at lower prices in mainland China and Gavi-eligible countries.^[46,47]^ For Furthermore, a new 9vHPV vaccine (Cecolin-9), which has recently been approved for marketing in mainland China and is expected to be marketed at a price 50% lower than the imported second-generation 9vHPV vaccine (Gardasil-9),^[48,49]^ could be a cost-effective alternative if it demonstrates noninferior clinical protection compared to Gardasil-9 in phase 3 trials.^[50]^

Considering the uncertainty of vaccine uptake in boys and the protection duration of the one-dose schedule, our analysis highlights the potential benefits and trade-offs of various vaccination strategies (summarized in Table 1). Under the base case vaccination cost, 1F1M was the most cost-effective strategy although the uncertainty around this conclusion increased if one-dose only gives 20 years of protection and boys’ uptake is low. However, 2F1M was always more beneficial than 2dFOV, and would be more cost-effective than 2dFOV in scenarios of 25%/50% uptake in boys. These findings suggest the potential of 1F1M vaccination while monitoring vaccine uptake among boys and duration of protection for one-dose vaccination from international studies such as the International Agency for Research on Cancer (IARC) India trials which currently showed no evidence of waning from one-dose vaccination in 12 years of follow-up.^[51]^ However, if the cost of vaccination is greatly lowered, then 2F1M could also be considered for better health benefits, particularly at high vaccine uptake in boys.

**Table 1.** The cost-effective vaccination strategy at the base case vaccination cost. ^a, b^.

| Duration of vaccine-induced protection by the one-dose schedule | Vaccine uptake among boys <sup>c</sup> |  |  |
| --- | --- | --- | --- |
|  | 25% | 50% | 85% |
| 20-year | 2F1M <sup>d</sup> | 1F1M | 1F1M |
| 30-year | 1F1M | 1F1M | 1F1M |

Abbreviations: FOV, female-only vaccination; GNV, gender-neutral vaccination; QALY, quality-adjusted life year; 1F1M, one-dose schedule for both schoolgirls and schoolboys; 2dFOV, two-dose schedule for FOV; 2F1M, two-dose schedule for schoolgirls and one-dose schedule for schoolboys.
<sup>a</sup> We considered a GNV strategy effective if it incurred more QALYs (mean and median) than the status quo 2dFOV.
<sup>b</sup> The findings were based on the base case scenario that assumed all HPV-related anal cancers in males are equally avertable by FOV via indirect protection and did not take into account of the impact of HPV vaccination on genital warts.
<sup>c</sup> Vaccine uptake among schoolgirls was assumed to be 85% in all strategies.
<sup>d</sup> The two-dose schedule was assumed to provide lifelong protection.

## Limitations

Our study has several limitations. First, there are no published local data on HPV prevalence and HPV type distribution for sites other than the cervix and oral cavity.^[14,40,52]^ As such, we used the HPV type distribution among HPV-positive cancer cases in Asia and Oceania in model calibration.^[2]^ Second, our statistical models that link genital HPV prevalence to the incidence of non-cervical HPV-related cancers did not include precancerous lesions such as anal or penile intraepithelial neoplasia.^[53]^ Without data for the intermediate precancerous lesions, the non-cervical HPV-related cancer model specification and calibration cannot be fully validated.^[54]^ Third, we did not explicitly include MSM in the model structure. Local data on the distribution of MSM among HPV-related cancer cases was unavailable. This limited our ability to perform a thorough evaluation of the effects of HPV vaccination in the MSM subgroup and the general population. To address this limitation, we ran a scenario analysis in which 40% of male anal cancers occurred among homosexual MSM, which was the same as assuming that FOV could avert at most 60% of HPV-related anal cancers in males.^[31]^ In this scenario, 2F2M remained not cost-effective regardless of vaccine uptake among boys.

## Conclusions

Our findings suggested that 2F2M was not cost-effective compared to 2dFOV unless the current vaccination cost is substantially reduced. 1F1M was likely cost-effective compared to 2dFOV but its health impact was sensitive to the protection duration of the one-dose schedule. 2F1M would be a cost-effective alternative to 2dFOV if vaccination cost could be moderately lowered and vaccine uptake of 85% in boys can be achieved.

## Data Availability

The data generating the findings of this article are included within the article and its additional file.

## Acknowledgements

This work was supported by grants from the Health and Medical Research Fund (CID-HKU-2-20) of the Government of the Hong Kong Special Administrative Region. This work was supported by AIR@InnoHK administered by the Innovation and Technology Commission. The computations were performed using research computing facilities offered by Information Technology Services, at the University of Hong Kong.

