## Supplementary file for "Cost-effectiveness and health impact of gender-neutral and single-dose HPV vaccination in Hong Kong"

**eFigures**

**eTables**

### S1. Methods

**S1.1. Overview of the *natural history model of cervical cancer* and *transmission dynamics* of HPV infection**

We expanded our previously developed model [1] to study the change in HPV infection by including boys in the current girls-only HPV vaccination program. Our previous model considered the natural history of HPV infection and the development of cervical cancer among females. For females, the natural history included health states of normal (no HPV infection), HPV infection, precancerous lesions (cervical intraepithelial neoplasia [CIN] with stages 1, 2, and 3), asymptomatic preclinical cervical cancer, and symptomatic clinically diagnosed cervical cancer. The natural history model for males included health states of normal and HPV infection. HPV infection rates are dependent on the age and sexual activity level of individuals, and the sexual mixing patterns between genders. The natural history models for both females and males also included a health state of immune which refers to HPV type-specific natural immunity after natural recovery of HPV infection. We considered four groups of high-risk oncogenic HPV types in the models: (a) HPV-16, (b) HPV-18, (c) HPV-OV which combines the other five vaccine-targeted high-risk HPV types (i.e., HPV-31, 33, 45, 52, and 58), and (d) HPV-NV which groups all other non-vaccine high-risk HPV types. Co-infection of multiple groups of HPV types is allowed in the model.

The model included a dynamic component to capture the transmission of HPV infection between heterosexual females and males in the population. The models stratified the population into 76 age cohorts (10, 11, …, 84, and 85+ years old) and three sexual activity levels for the number of sexual partners during the past six months (none, low, and high) for both female and male populations. We used the sexuality survey data reported by the Family Planning Association of Hong Kong (FPAHK) to construct the sexual contact patterns of both genders.[2] We assumed that the population was sexually naïve before the age of 10 years. Individuals may be infected with HPV via sexual activities when they become sexually active at older ages. The transmission dynamic component simulated the heterosexual transmission of HPV infection between males and females before and after HPV vaccination. The dynamic component captured herd immunity in the entire population, i.e., the indirect protection to unvaccinated individuals due to the immunity in the population generated by vaccinees.

In model calibration, we estimated (i) natural history parameters relating to HPV type-specific transmissibility and the development of cervical cancer and precancerous lesions (e.g., transition rates between various health states), and (ii) sexual mixing parameters including the degree of assortativeness across ages and sexual activity levels, and the spread of age preference in forming sexual partnerships. We used Markov chain Monte Carlo (MCMC) to infer these parameters by fitting the model outcomes to empirical data including local age- and HPV type-specific genital HPV prevalence among females, and age-specific cervical cancer incidence. The uncertainty of the inferred parameters was taken into account in the health economic evaluation by considering the parameter sets from the posterior distribution in the probabilistic sensitivity analysis (PSA).

**S1.2. Modeling the incidence of *non-cervical HPV-related cancers***

*Linking the incidence of non-cervical HPV-related cancers with HPV incidence from the transmission dynamic model*

We modeled the incidence of non-cervical HPV-related cancers based on the genital HPV incidence rate of the respective genders.[3,4] eFigure 1 shows the schematic presentation. We first obtained the age- and HPV type-specific genital HPV incidence rate for each gender from the transmission dynamic model for cervical cancers (i.e., the model described in S1.1).[1] For each non-cervical HPV-related cancer, we linked the age- and HPV type-specific genital HPV incidence to age-specific incidence of cancer with an age-specific scaling function and a time-lagged function. Referring to the data reported in the Hong Kong Cancer Registry (HKCaR),[5] we fitted the modeled cancer incidence with empirical cancer incidence at 5-year age groups of 25-29, 30-34, …, 80-84, and 85+ years. We also fitted the outcomes to the HPV type distribution in HPV-positive cancer cases reported in the literature.[6,7] In this study, we considered oropharyngeal (OPC), anal, penile, and vaginal/vulvar cancers for non-cervical HPV-related cancers. In the HKCaR database, penis and other male genital cancers are grouped in one category. Similarly, a single categorization is used for vagina/vulva and other female genital cancers. We estimated the age-specific incidence of anal cancers based on the proportion of cancers at the anus in the rectum.[5,8]

Without loss of generality, the below notations are applied to each non-cervical HPV-related cancer for either gender and each HPV type $q$ unless otherwise specified.
Let $C^q(a)$ be the age-specific cancer incidence at age $a$,
$d(t)$ be a time-lagged function,
$s(a)$ be an age-specific scaling function, and
$h(a)$ be the age-specific genital HPV incidence at age $a$ obtained from the transmission dynamic model.
Here, $d(t)$, $s(a)$, and $h(a)$ are specific to HPV type $q$ (but we suppress the notation $q$ in the formula for simplicity). Then, the age-specific cancer incidence at a non-cervical site for HPV type $q$ can be expressed as:

$$C^q(a) = \int_0^\infty s(a-t)h(a-t)d(t)dt\,. \qquad \ldots \quad (1)$$

Following the above, we modeled $C^q(a)$ for non-cervical site-specific cancer incidence for HPV type $q$ at age $a$. We linked $C^q(a)$ with $h(a-t)$ for the genital HPV incidence at earlier ages which was obtained from the transmission dynamic model. We modeled $C^q(a)$ regardless of the attributable fraction of HPV infection among cancer cases. The attributable fraction of HPV infection in non-cervical cancers would be taken into account when evaluating the changes in cancer incidence following large-scale HPV vaccination (please refer to S1.3 for details).

For parameter inference for non-cervical HPV-related cancer, we estimated parameters for (a) two scaling factors at ages 10 and 40 years for $s(a)$ for the age-specific scaling function at age group $a$, and (b) the distribution of the time-lagged function $d$, i.e., the scale and shape parameters for assuming a gamma distribution for the lag time. We assumed that the scaling factor $s(a)$ increases linearly from age 10 to 40 years, and then becomes constant after age 40 years. We estimated HPV type-specific parameters for HPV types HPV-16, HPV-18/OV, and HPV-NV.

*Likelihood of the fitting*
<u>Fitting to cancer incidence</u>
Let $C(a)$ be the overall age-specific cancer incidence rate among different HPV types at age group $a$ based on the model, and
$X(a)$ be the observed age-specific cancer incidence rate at age group $a$ based on HKCaR.[5]

Formula (1) presents an age-specific cancer incidence for each of the three HPV types $q$ (16, 18/OV, and NV). The sum of the age- and HPV type-specific incidences forms the overall age-specific cancer incidence $C(a)$ (i.e., $C(a) = \sum_q C^q(a)$). The modeled overall cancer incidence $C(a)$ was compared to the observed incidence statistics $X(a)$.

Let $logL_C$ be the log-likelihood of the fitting on cancer incidence. Assuming that the cancer incidence follows a Poisson distribution, then
$logL_C = \sum_{j\in a}\big(X(j)\log\big(C(j)\big) - C(j) - \log(X(j)!)\big)$, where $j$ refers to the age groups.

<u>Fitting to HPV relative prevalence in HPV-positive cancer cases</u>
Based on the calculation of cancer incidence for each HPV type, we obtained the modeled proportions of each HPV type among the overall cancer cases.

Let $p_{16}^M, p_{18/OV}^M, p_{NV}^M$ be the modeled proportions of cancer cases across ages for each HPV type;

$n$ be the reported number of cancer samples that were HPV-positive in clinical studies; and $n_{16}, n_{18/OV}, n_{NV}$ be the reported numbers of cancer samples that were tested positive for each HPV type HPV-16, 18/OV, and NV in the clinical studies, respectively.

We referred to Chen et al. (2024) for the relative prevalence of HPV types in OPC in the Hong Kong setting.[7] We considered the relative prevalence of HPV types in Asian & Oceanian studies reported in the meta-analysis by de Sanjose et al. (2019) for other non-cervical HPV-related cancers given the lack of corresponding data reported in Hong Kong.[6]

Let $logL_H$ be the log-likelihood of the fitting on the relative prevalence of high-risk HPV types among HPV-positive cancer cases. We assumed that the relative prevalence followed a multinomial distribution. Then, $logL_H = \log(n!) - \sum_q \log(n_q!) + \sum_q (n_q \times \log(p_q^M))$, where $q$ refers to HPV types HPV-16, 18/OV, and NV.

The overall log-likelihood would be $logL = logL_C + logL_H$.

We inferred the parameter set of (a) the scaling factor for each HPV type/group, and (b) the distribution of the lag time using MCMC. We estimated parameter sets for each non-cervical HPV-related cancer and each gender separately. We used the modeled age-specific genital HPV incidence based on the posterior distribution from our transmission dynamic model as the input $h(a)$ in the above formula.[1]

eFigure 2 presents the calibration for comparing the modeled and empirically observed age-specific non-cervical HPV-related cancers. eFigure 3 presents the comparison of the relative prevalence of HPV-16 and HPV-16/18/31/33/45/52/58 (HR-9vHPV; high-risk HPV types that are covered by the nonavalent (9vHPV) vaccine) among HPV-positive cases for each non-cervical HPV-related cancer.

**eFigure 1**. Schematic presentation of modeling incidence of cervical cancer and non-cervical HPV-related cancers.

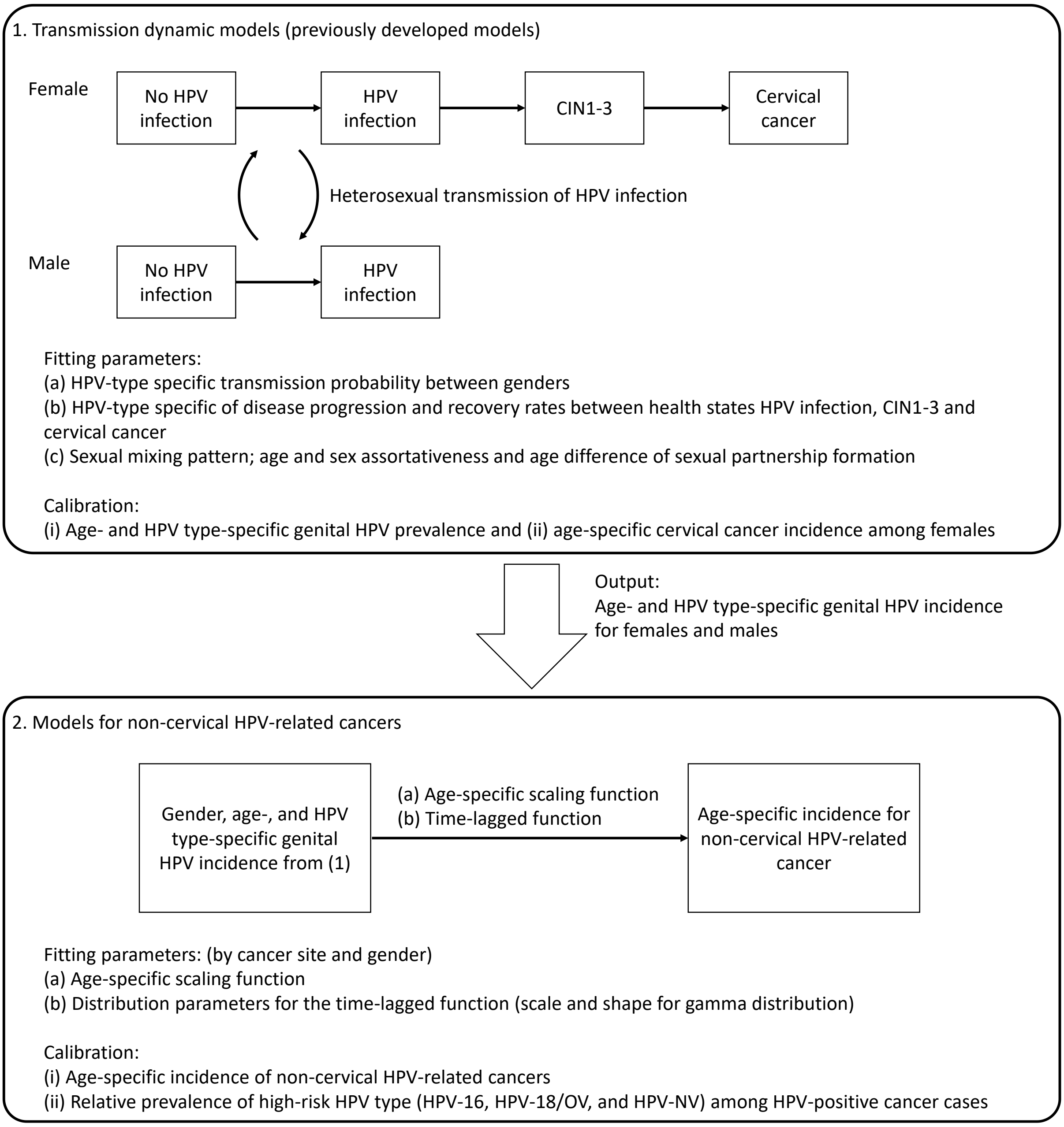

Abbreviations. CIN, cervical intraepithelial neoplasia; NV, non-vaccine-targeted high-risk HPV types (i.e., high-risk HPV types other than HPV-16/18/31/33/45/52/58); OV, high-risk HPV types that were targeted by 9vHPV vaccines other than HPV-16/18 (i.e., HPV-31/33/45/52/58).

Notes. [(a)] The transmission dynamic model was developed and calibrated to age- and type-specific HPV prevalence and age-specific cervical cancer incidence rate.[1] HPV type-specific disease progression rates, recovery rates, and sexual mixing parameters were inferred during model calibration. [(b)] Gender-, age-, and HPV type-specific HPV incidence from model 1 was input in model 2 for modeling the incidence of non-cervical HPV-related cancers. For each non-cervical HPV-related cancer, we estimated (a) the age-specific scaling function, and (b) the distribution parameters for the time-lagged function. HPV type-specific parameters for HPV types 16, 18/OV, and NV were inferred. We considered (i) the observed overall age-specific incidence of non-cervical HPV-related cancers, and (ii) the relative prevalence of high-risk HPV types among HPV-positive cancer cases as the fitting targets in model calibration.

**eFigure 2**. Model calibration of age-specific incidence of non-cervical HPV-related cancers.

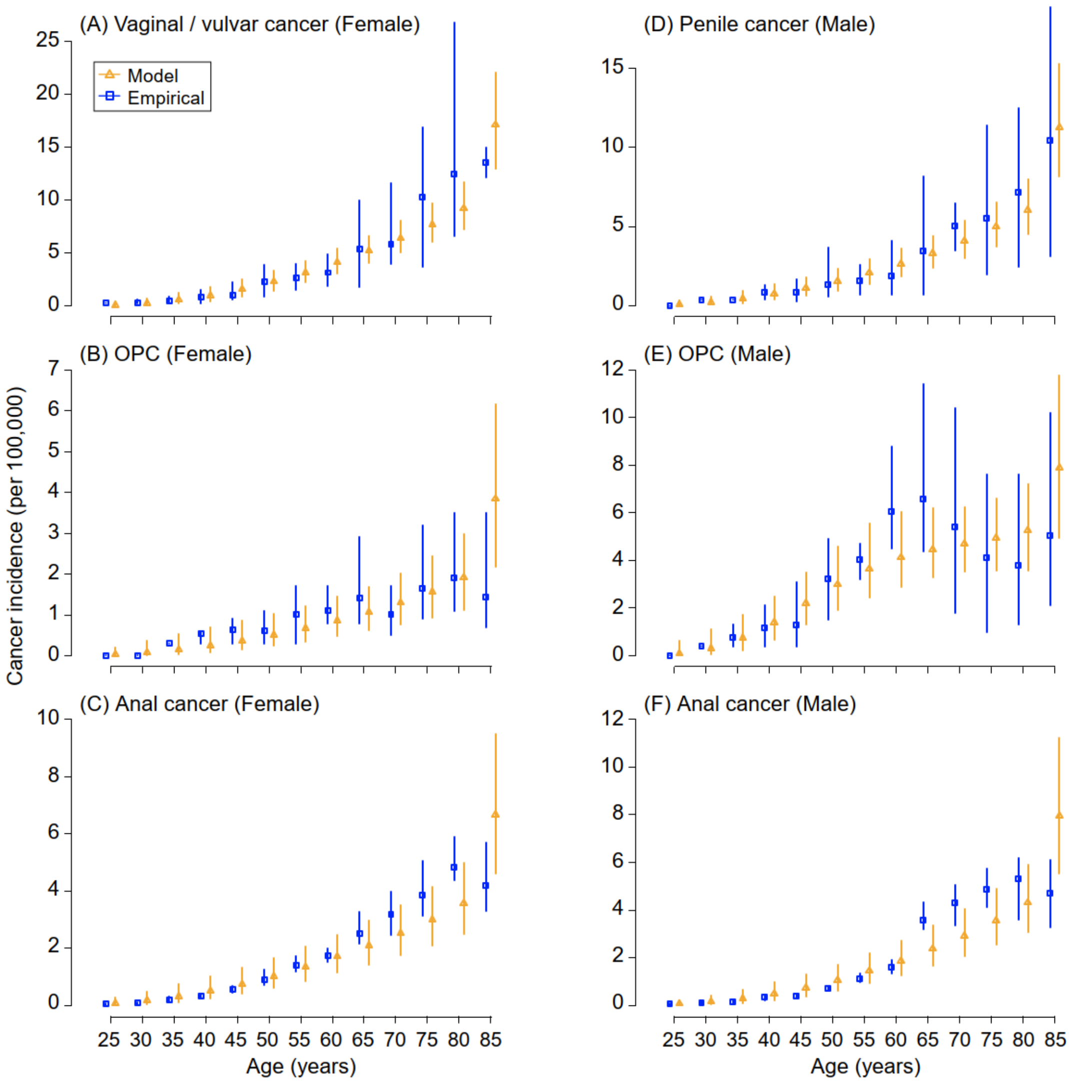

Notes. Each plot presents the comparison of the empirical incidence from the Hong Kong Cancer Registry (blue squares and lines)[5] and the modeled incidence from calibration (orange triangles and lines) for non-cervical HPV-related cancer. The mean and the range of the corresponding cancer incidence reported in 2012-2021 were presented for the empirical incidence (blue squares and lines, respectively). The posterior medians and 95% credible intervals were presented for the modeled incidence (orange triangles and lines, respectively).

**eFigure 3**. Comparison between modeled and empirical relative prevalence of high-risk HPV types in HPV-positive non-cervical cancers in (A) females and (B) males.

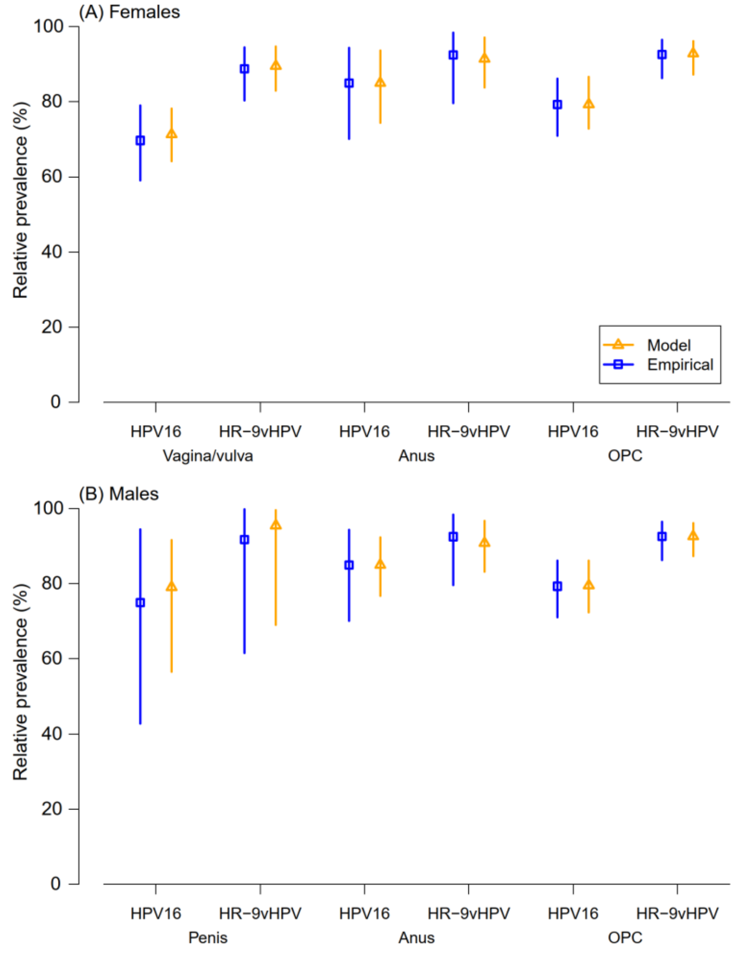

Notes. [(a)] Relative prevalence refers to the proportion of the respective HPV type/class among HPV-positive cancer cases. For the empirical relative prevalence, we considered the findings in de Sanjose et al (2019) for vaginal/vulvar, penile, and anal cancers based on the Asia & Oceania subgroup,[6] and Chen et al (2024) for OPC based on a Hong Kong study.[7] [(b)] HR-9vHPV refers to high-risk HPV types 16/18/31/33/45/52/58 that are covered by the 9vHPV vaccines. [(c)] Blue squares and lines present the point estimates and 95% confidence intervals for proportions of relative prevalence retrieved from the literature, respectively. Orange triangles and lines present the posterior medians and 95% credible intervals for proportions generated from model calibration, respectively.

**S1.3. Estimating the change in non-cervical HPV-related cancer incidence after large-scale HPV vaccination**

To estimate the change in non-cervical HPV-related cancer incidence after large-scale HPV vaccination, we adopted and modified the approach that Jit et al used for evaluating the impact of female and male HPV vaccination in the English setting.[9] For simplicity, we suppress the notation of the particular site $S$ for a non-cervical HPV-related cancer in the following formula. Let

$N(a,t)$ be the annual incidence of a non-cervical HPV-related cancer of a particular site $S$ at age group $a$ and $t$ years after HPV vaccination;

$C_0(a)$ be the annual incidence of a non-cervical HPV-related cancer of a particular site $S$ at age group $a$ prior to HPV vaccination;

$C^M(a,t)$ be the modeled annual incidence of a non-cervical HPV-related cancer of a particular site $S$ at age group $a$ and $t$ years after HPV vaccination, and be estimated based on the HPV incidence at $t$ years after HPV vaccination by incorporating the site- and gender-specific parameters inferred in S1.2;

$x$ be the attributable fraction for the proportion of cancer in site $S$ that is attributable to HPV infection.

We assumed that (i) the incidence of cancer cases that are attributable to HPV infection (i.e., $x$) would be changed following HPV vaccination, and (ii) the incidence of cancer cases that are not attributable to HPV infection (i.e., $(1-x)$) would not be changed by HPV vaccination.

Then, the age-specific annual incidence of a non-cervical HPV-related cancer of a particular site $S$ at $t$ years after vaccination, $N(a,t)$, can be estimated with the following formula:

$$N(a,t) = x \times C^M(a,t) + (1-x) \times C_0(a) \; .$$

For the proportion of non-cervical HPV-related cancer that is attributable to HPV infection (i.e., $x$), we referred to local literature when relevant data is available, or otherwise regional or global studies (eTable 1).[7,10-13] We considered the uncertainty of the attributable fraction of HPV infection in PSA.

**eTable 1**. Attributable fraction for the proportion of non-cervical HPV-related cancers that are attributable to HPV infection and the uncertainty distribution used in the probabilistic sensitivity analysis.

| Gender | Cancer site | Mean | Lower 95% CI | Upper 95% CI | Beta *a* [a] | Beta *b* [a] | Reference |
|---|---|---|---|---|---|---|---|
| Female | Oropharynx/Tonsil | 0.419 | 0.348 | 0.492 | 75 | 104 | [7] |
| Female | Vagina | 0.630 | 0.488 | 0.762 | 29 | 17 | [10] |
| Female | Vulva | 0.211 | 0.158 | 0.276 | 40 | 148 | [11] |
| Female | Anus | 0.854 | 0.814 | 0.890 | 281 | 48 | [12] |
| Male | Oropharynx/Tonsil | 0.419 | 0.348 | 0.492 | 75 | 104 | [7] |
| Male | Penis | 0.104 | 0.044 | 0.187 | 7 | 60 | [13] |
| Male | Anus | 0.803 | 0.737 | 0.861 | 126 | 31 | [12] |

Abbreviation: CI, confidence interval.

Notes. [a] Beta *a* and Beta *b* refer to the *alpha* and *beta* parameters in a Beta distribution for probabilistic sensitivity analysis, respectively.

### S2. Cost-effectiveness analysis

*HPV vaccination*

Under the current female-only HPV vaccination (FOV) immunization program in Hong Kong, primary five and six schoolgirls could receive two doses of 9vHPV vaccines for free via school-based immunization service or at the sub-offices of the immunization team by appointment.[14] We assumed that the same arrangement for both eligible girls and boys would be used for the implementation of gender-neutral vaccination (GNV). We considered three scenarios for vaccine uptake among schoolboys in GNV: (i) a high uptake of 85% as observed among schoolgirls, (ii) a moderate uptake of 50% based on a local survey conducted in 2016 on parental acceptability of free HPV vaccination for boys aged 9-13 years,[15] and (iii) a low uptake at 25% based on another local survey on male baccalaureate students aged 18-26 years.[16] We assumed that expanding the program from the status quo two-dose FOV (2dFOV) to GNV would not affect vaccine uptake among girls.

Regarding vaccine efficacy, we assumed that the 9vHPV vaccines provide the same efficacy to vaccine-targeted HPV types to both genders. Specifically, we set the vaccine efficacy at 95.5% (95% CI 90.0% to 98.4%) against HPV-16, 95.8% (84.1% to 99.5%) against HPV-18, 96.0% (94.4% to 97.2%) against HPV-OV (HPV-31, 33, 45, 52, and 58).[1,17,18] We set 0% efficacy in preventing non-vaccine HPV types. We assumed that the two-dose vaccination schedule provided vaccinees lifelong protection.[19] For the one-dose vaccination schedule, based on the latest findings of clinical trials with an average follow-up of 12 to 16 years,[20,21] we assumed a fixed duration of 20- or 30-year vaccine-induced protection to vaccinees.[19,22] We assumed that the one-dose and two-dose vaccination schedules provide the same vaccine efficacy against vaccine-targeted HPV types.[20,21] We considered two-dose and one-dose GNV strategies (2F2M and 1F1M, respectively) that schoolgirls and schoolboys received the same number of doses of HPV vaccines. We also explored the potential of 2F1M for adding a single dose for boys on top of the status quo 2-dose FOV schedule for girls.

*Cervical screening*

The cervical screening program in Hong Kong recommends that females aged 25-64 years attend screening for cervical cancer with a routine screening interval of every 3 or 5 years following negative cytology or HPV testing, respectively.[23] We assumed that the cervical screening recommendation and screening attendance would be the same in the era of HPV vaccination. Following the latest practice issued by the Department of Health in April 2023, we considered using HPV testing as the primary screening modality for women aged 30-64 years and cytology for women aged 25-29 years.[24] We assumed that 70% of females aged 25-64 years would attend cervical screening.[25] We followed the history-dependent screening and management guidelines issued by the Hong Kong College of Obstetricians and Gynecologists (HKCOG).[26] Females who are diagnosed with CIN2/3 and preclinical cervical cancer will be treated accordingly. We assumed that screening uptake would remain unchanged (i.e., at 70%) when assessing the cost-effectiveness of vaccination strategies.

*Cost-effectiveness analysis*

We conducted a cost-effectiveness analysis of expanding the current HPV vaccination program from girls-only to including both girls and boys using a societal perspective. We considered the costs of cancer treatment and cervical screening costs from the corresponding private charges stated in the Government's Gazette by the Hospital Authority.[27] The private charges exclude subsidies from the government. We assumed that public hospitals make these charges for covering their costs instead of making a profit. The Hospital Authority covers approximately 90% of hospitalization services in Hong Kong,[28] so the charges stated in the Gazette provide reliable public health estimates. Females' time cost and transportation cost for attending cervical screenings or clinical appointments were included. We set the base case vaccination cost at US$218 (HK$1,700) per dose (including vaccine cost and administration expenses) for both genders by referring to the corresponding price listed by the FPAHK.[29] FPAHK is a non-profit making organization providing sexual and reproductive health services for the community in Hong Kong. We assumed that the charge of vaccination listed by FPAHK covers the cost instead of making a profit. We quantified the health benefits by quality-adjusted life years (QALYs). Given the lack of local estimates on health utilities, we used the health utilities that were used in international and local CEAs on cervical cancers and cervical screening as well as other non-cervical HPV-related cancers.[9,30,31] Furthermore, stage distribution and relative survival are not available for non-cervical HPV-related cancers in Hong Kong. We referred to the stage distribution and stage-specific 5-year relative survival for each non-cervical HPV-related cancer reported in the Surveillance, Epidemiology, and End Results (SEER) Program in the United States when calculating the average treatment costs and QALYs for each cancer case for non-cervical HPV-related cancers.[32]

We considered the incremental cost-effectiveness ratio (ICER) as the main outcome measure in the study. ICER was defined as the incremental cost divided by the incremental QALYs gained for expanding FOV to GNV. We considered one gross domestic product per capita (GDPpc) for each QALY gained as the WTP threshold, as done in previous health technology assessments of a similar nature in Hong Kong.[1,8] The average local GDPpc was US$48,757 (HK$380,303; US$1 = HK$7.8) in the recent 5 years (2019-2023).[33] We also referred to the findings of a recent study which estimated the cost-effectiveness thresholds based on the national health expenditures per capita and life expectancy for 174 countries.[34] From that global study, among countries that have a similar life expectancy (>80 years) and GDPpc (ranging from US$45,000 to US$55,000) to Hong Kong, the mean of the estimated cost-effectiveness threshold was US$46,181, which is similar to the WTP threshold we used. Furthermore, while we considered three scenarios on vaccine uptake among boys (i.e., 25%, 50%, and 85%), because vaccine uptake is not a decision variable, we did not evaluate the ICER across different uptake scenarios.

We estimated the threshold vaccination cost (TVC) for expanding FOV to GNV. TVC per dose was defined as the vaccination cost that includes vaccine cost and administration expenses for each vaccine dose. When vaccination cost equals TVC, the intervention and the comparator would be equally cost-effective at the WTP threshold. Based on the TVC, we calculated the corresponding relative change and relative reduction compared to the base case vaccination cost (US$218 per dose). The relative change is expressed as $\left(\frac{\text{TVC}}{\text{VaccinationCost}_{\text{basecase}}}\right) \times 100\%$. The relative reduction is expressed with the formula: $\left(1 - \frac{\text{TVC}}{\text{VaccinationCost}_{\text{basecase}}}\right) \times 100\%$. A positive relative reduction means that the estimated TVC was lower than the base case vaccination cost. That is, the vaccination cost has to be reduced for the intervention and comparator to be equally cost-effective. A negative relative reduction of a TVC refers to the relative increase in the base case vaccination cost if GNV was cost-effective compared to 2dFOV with an ICER below the WTP threshold at the base case vaccination cost. In this case, a higher vaccination cost would be acceptable for the intervention and comparator to be equally cost-effective at the WTP threshold. For example, a relative reduction of $-100\%$ means that the TVC would be a relative increase of 100% in the base case vaccination cost, i.e., the TVC is double the base case vaccination cost. We applied the same mechanism when comparing the cost-effectiveness of 2F1M to 1F1M.

We accounted for the uncertainty of input parameters in the PSA. We sampled 100 sets of disease-related parameters about (i) disease transition rates for the development of cervical cancer and precancerous lesions (e.g., progression rates from HPV infection to CIN1, from CIN1 to CIN2 and from CIN2 to CIN3), (ii) sexual mixing pattern (e.g., the degree of assortativeness across ages and sexual activity levels, and the spread of age preference in forming sexual partnerships), (iii) statistical linkages between genital HPV infection and incidence of non-cervical HPV-related cancers, and (iv) vaccine efficacies. We also sampled 100 parameter sets about the costs of screening and treatment and health utilities. That is, a total of 10,000 combinations were considered in the PSA. eTable 2 presents the cost and the health utility parameters that were used in the analysis.

*Scenario analysis*
We conducted sensitivity analyses to account for the disease burdens associated with anal cancers in males, and genital warts in males and females. Regarding anal cancers in males, we assumed in the base case that all HPV-related anal cancers in males are equally avertable by FOV via indirect protection. In a sensitivity analysis, we considered an alternative scenario in which only 60% of HPV-related anal cancers in males were avertable by FOV, based on the proportion (40%) of male anal cancer patients who were men who have sex with men (MSM) in the United States.[35]

Regarding genital warts, in the base case, we focused on the impacts of HPV vaccines on HPV-related cancers when evaluating the impacts of implementing GNV and did not take into account the cases of genital warts. In a sensitivity analysis, we considered a scenario that accounted for the costs and health associated with genital warts. We assumed that 90% of genital warts were attributable to infection by low-risk HPV types 6/11,[36] which are preventable by the 9vHPV vaccines. A local study estimated that the incidence rate of genital warts was 292.19 in men and 124.86 in women per 100,000 person-years, based on new cases of genital warts diagnosed from patients in both public and private primary care clinics in January 2009.[37] We projected and estimated the change in the incidence of genital warts due to HPV-6/11 after HPV vaccination by assuming that HPV 6/11 incidence reduced in proportion to the reduction in HPV-16/18 incidence estimated from our transmission dynamic model.

**eTable 2**. Cost and health utility parameters that are used in the analysis.

| Cost items | Distribution (US$) [a] | References |
|---|---|---|
| Cytology test | *T*(37.2, 111.5, 74.4) | [27] |
| HPV test | *T*(65.4, 130.8, 87.2) | [27] |
| Colposcopy + biopsy | *T*(552.6, 1157.7, 855.1) | [27] |
| Treatment for CIN2 or CIN3<br>Loop electro-surgical excision procedure (LEEP) | *N*(2164.1, 541.0) | [27] |
| Treatment for Stage I cervical cancer<br>Wertheim's hysterectomy | *N*(15,510.3, 3877.6) | [27] |
| Treatment for Stage II-III cervical cancer<br>Radiotherapy + chemotherapy + brachytherapy | *N*(48,212.2, 12,053.1) | [27] |
| Treatment for Stage IV cervical cancer<br>Radiotherapy + chemotherapy | *N*(24,902.3, 6225.6) | [27] |
| Palliative care hospitalization (per day) | *T*(567.9, 852.6, 710.3) | [27] |
| Staff cost for screening | *N*(25.6, 6.4) | [27] |
| Staff cost for treatment of precancerous lesions | *N*(109.0, 27.3) | [27] |
| Time cost (half day) | *N*(31.3, 7.8) | [38] |
| Transportation | *N*(6.4, 1.6) | [39,40] |
| Treatment cost for non-cervical HPV-related cancers | | [8] |
| Anal cancer | *N*(31,614.7, 7,903.7) | |
| Oropharyngeal cancer (OPC) | *N*(35,652.3, 8,913.1) | |
| Penile cancer | *N*(24,843.6, 6,210.9) | |
| Vaginal/vulvar cancer | *N*(19,645.7, 4,911.4) | |
| Treatment cost for genital warts | 141 | [8] |
| **Health outcomes** | **Distribution** | **References** |
| Utility loss per episode of screen results | | |
| Negative cytology / negative HPV test | *T*(0.00002, 0.00023, 0.0001) | [31] |
| ASCUS | *T*(0.00023, 0.002, 0.0011) | [31] |
| Positive HPV test | *T*(0.00023, 0.0089, 0.004) | [31] |
| Normal colposcopy | *T*(0.0015, 0.04, 0.0147) | [31] |
| LSIL / CIN1 | *T*(0.005, 0.11, 0.0618) | [31] |
| CIN23 | *T*(0.003, 0.13, 0.0783) | [31] |
| Health utility during and post-treatment of cancers | | |
| During treatment (6 months or until death) | | |
| Stage I | *T*(0.49, 0.81, 0.705) | [31,41] |
| Stage II | *T*(0.42, 0.67, 0.615) | [31,41] |
| Stage III | *T*(0.42, 0.70, 0.56) | [41] |
| Stage IV | *T*(0.36, 0.60, 0.48) | [41] |
| Post-treatment (4.5 years or until death) | | |
| Stage I | *T*(0.73, 0.99, 0.97) | [31,41] |

| | | |
|---|---|---|
| Stage II | $T$(0.68, 0.98, 0.935) | [31,41] |
| Stage III | $T$(0.68, 0.98, 0.935) | [31,41] |
| Stage IV | $T$(0.47, 0.969, 0.795) | [31,41] |
| Utility loss per episode of genital warts | 0.018 | [42] |

Notes. [(a)] $N(a, b)$ denotes a normal distribution with mean $a$ and standard deviation $b$. A coefficient of variation of 0.25 was considered when the standard deviation was unavailable for the normal distribution. $T(a, b, c)$ denotes a triangular distribution that ranges from $a$ to $b$ with mode $c$.

### S3. Additional findings on the cost-effectiveness and health impacts of GNV strategies

This section presents the additional findings regarding the CEA and health impacts for comparing GNV vs FOV.

#### S3.1. Findings on CEA

*1F1M*

eFigure 4 presents the cost-effectiveness planes for 1F1M vs 2dFOV for the base case and alternative scenarios if the one-dose schedule provided 20-year protection to vaccinees. In the base case scenario, if the vaccine uptake among boys was 25% and 50%, 1F1M resulted in fewer QALYs than 2dFOV in 75% and 33% of the simulations, respectively (eFigure 4A). However, if the one-dose schedule provided 30-year protection to vaccinees, 1F1M generated more QALYs in most simulations when compared to 2dFOV (eFigure 5). In the base case scenario, compared to 2dFOV, if vaccine uptake among boys was 25% and 50%, 1F1M was cost-saving with more QALYs in 91% of and all simulations, respectively (eFigure 5A).

1F1M may incur fewer QALYs compared to 2dFOV, depending on the protection duration of the one-dose schedule and vaccine uptake among boys (assuming that vaccine uptake among girls remained unchanged). A high assortativeness by age in the formation of sexual partners (i.e., the sexual mixing) and a high relative HPV transmission probability at an older age were the two main model parameters contributing to fewer QALY gained. A high age assortativeness in sexual mixing responds to vaccination impact being more concentrated in a narrow age range of vaccinees. A high relative HPV transmission probability at older ages means that HPV transmission would still be high at older ages (please refer to our previous work in Choi 2018[1] for the definition of the relative HPV transmission probability in the natural history model). In both cases, a greater vaccine impact, such as if vaccine uptake was higher or vaccines provided longer protection, would be needed to lower HPV transmission across genders.

*2F1M*

eFigure 6 presents the ICERs and TVCs for comparing 2F1M to 2dFOV if the one-dose schedule was assumed to provide 30-year protection to vaccinees. The cost-effectiveness outcomes were similar to the comparison if the one-dose schedule provided 20-year protection (**Error! Reference source not found.** in the main text).

eFigure 7 and eFigure 8 present the ICERs and TVCs for comparing 2F1M to 1F1M, assuming that the one-dose schedule provided 20-year and 30-year protection to vaccinees, respectively. 2F1M was not a cost-effective alternative to 1F1M given the high ICERs and low TVCs.

*Distribution of treatment-related cost and QALY gained*

eFigure 9 presents the distribution of the difference in cost and QALY gained that were associated with treatment to the health outcomes of HPV-related cancers and genital warts for 2F2M vs 2dFOV for the base case scenario. The plots were prepared based on the median of the cost and QALY gained among PSA simulations. eFigure 10 presents the medians and 90% prediction intervals (PIs; 5$^{th}$ to 95$^{th}$ percentiles) of the difference in cost and QALY gained that were associated with treatment for HPV-related cancers and genital warts for comparing 2F2M to 2dFOV. eFigure 11 and eFigure 12 present the corresponding findings for 1F1M vs 2dFOV for the scenario if the one-dose schedule provided 20-year and 30-year protection, respectively. The findings suggest that when compared to 2dFOV, the greatest proportion of health impacts due to GNV are attributable to cervical cancer prevention, followed by male HPV-related cancers prevention (OPC and anal cancer) and genital warts.

*Numerical statistics for the cost-effectiveness of GNV strategies*

eTable 3 presents the ICERs, TVCs, and the percentage of simulations that were cost-effective at the WTP threshold and the base case vaccination cost. The table also presents the relative reduction of vaccination cost needed for 80%-95% of simulations to be cost-effective at the WTP threshold of one GDPpc when comparing 2F2M to the status quo 2dFOV, including the case for 90% of simulations to be cost-effective as listed by the Joint Committee on Vaccination and Immunisation (JCVI) in the United Kingdom for assessing immunization programs.[43] eTable 4 and eTable 5 provide the corresponding outcomes for 2F1M vs 2dFOV if the one-dose schedule was assumed to provide 20-year and 30-year protection, respectively. eTable 6 and eTable 7 present the outcomes for 2F1M vs 1F1M for the case if the one-dose schedule provided 20 years and 30 years of protection, respectively.

eTable 8 summarizes the cost-effectiveness of vaccination strategy for a sensitivity analysis that considers a GNV strategy effective if it incurred more QALYs than the status quo 2dFOV in at least 80% of simulations, i.e., a more risk aversion scenario compared to the setting in **Error! Reference source not found.** of the main text. In this setting, 1F1M remains the more cost-effective strategy in most scenarios. If the one-dose schedule provided 20 years of protection and vaccine uptake in boys was 50%, 1F1M resulted in more QALYs in 67% of simulations when compared to 2dFOV, suggesting that 2dFOV should be retained (instead of considering 1F1M as seen in the setting in Table 1 of the main text). In this scenario, 2F1M would be more cost-effective than 2dFOV if the vaccination cost could be reduced by 19%.

**S3.2. Findings on health impacts**

*HPV incidence of one-dose vaccination of shorter protection.*

eFigure 13 and eFigure 14 present the modelled HPV-16 infection among females and males, respectively. Compared to 2dFOV with lifelong protection, if changing to 1F1M of shorter protection (red and blue lines for 20 and 30 years of protection, respectively), a rebound of HPV-16 infection is seen in older ages when vaccine-induced protection wanes. The rebound was less apparent in incidence among males when compared to that among females. It was because GNV provided direct protection to males while FOV only provided them with indirect protection. A higher vaccine uptake in boys in 1F1M also delayed the rebound when compared to the scenarios of a lower boys' uptake. Specifically, if the one-dose schedule provided 20 years of protection, HPV-16 infection among males under 1F1M remained lower than that under 2dFOV until age 50 years if boys' uptake was 85% (eFigure 14C). However, the HPV infection in males with 1F1M implemented rebounded at age 40 years if uptake in boys was only 25% (eFigure 14A). On the other hand, compared to 2dFOV, HPV-16 infection among females under 1F1M rebounded at age 35 years regardless of vaccine uptake in boys if the one-dose schedule provided 20-year protection (eFigure 13). However, if the one-dose schedule provided 30-year protection, the rebound of female HPV-16 infection is seen at ages 45 and 50 years if uptake in boys was 25% and 85%, respectively, i.e., in a later age as vaccine uptake in boys increased.

*Reduction of genital warts*

eTable 9 presents the additional relative reduction of genital warts for comparing different GNV strategies to 2dFOV. We considered the overall cases of genital warts among age cohorts that were involved in the vaccination program over the time horizon of 100 years. Greater additional relative reductions of genital warts were observed in males than in females. It is because when compared to 2dFOV, including boys in GNV provided direct vaccine-induced protection to them, while FOV only provided them with indirect protection. For example, compared to 2dFOV, 2F2M prevented an additional 8.4% (3.4%, 16.1%) and 3.2% (1.2%, 6.7%) of genital warts in males and females, respectively, if vaccine uptake among boys is 85%. On the other hand, for 1F1M vs 2-dose FOV, if the one-dose and two-dose schedule was assumed to provide 20-year and lifelong protection, respectively, a negative additional relative reduction refers to the situation that 1F1M prevented fewer genital warts than 2-dose FOV, suggested by the rebound in HPV infection when shifting from 2dFOV to 1F1M with 20-year protection.

**eFigure 4**. Cost-effectiveness planes for 1F1M vs 2dFOV by vaccine uptake among boys (columns) in the base case and alternative scenarios (rows) if the one-dose schedule provided 20 years of protection.

Fewer simulations — More simulations; WTP threshold of 1 GDPpc; Mean of difference in discounted cost/QALY

(A) Base case

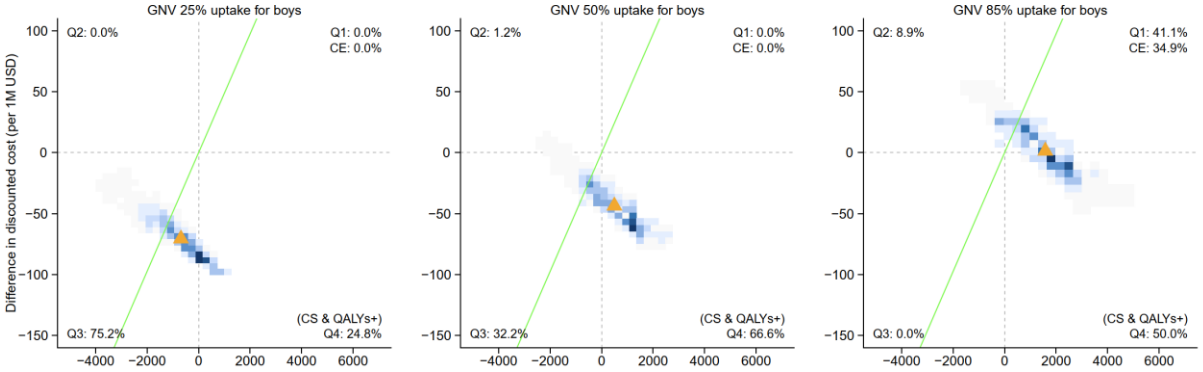

(B) 60% of HPV-related anal cancers in males were averted by FOV

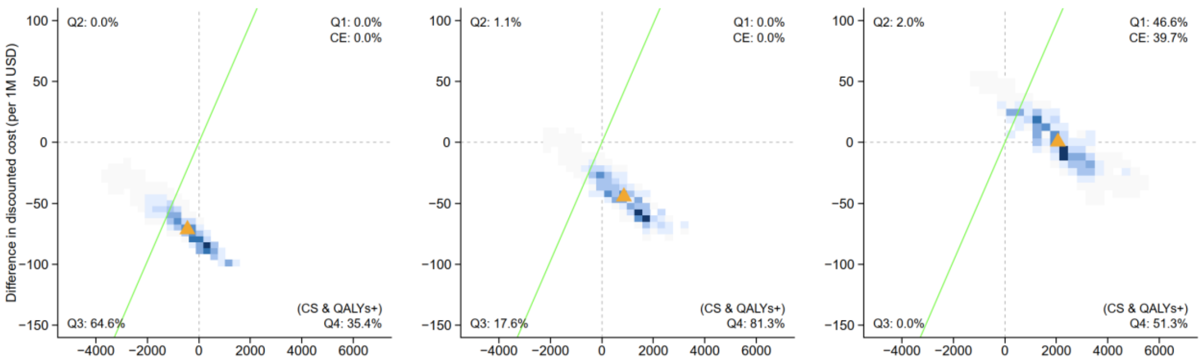

(C) Accounted for costs and QALYs associated with genital warts

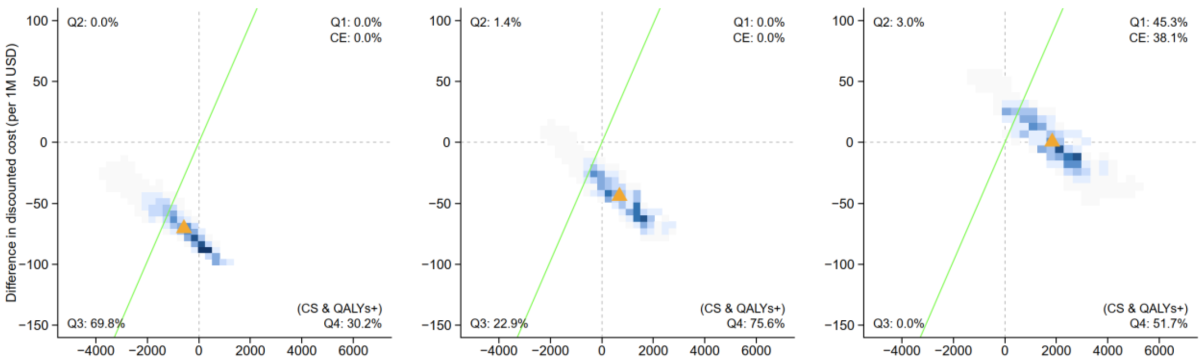

(D) Accounted for costs and QALYs associated with genital warts and 60% of HPV-related anal cancers in males were averted by FOV

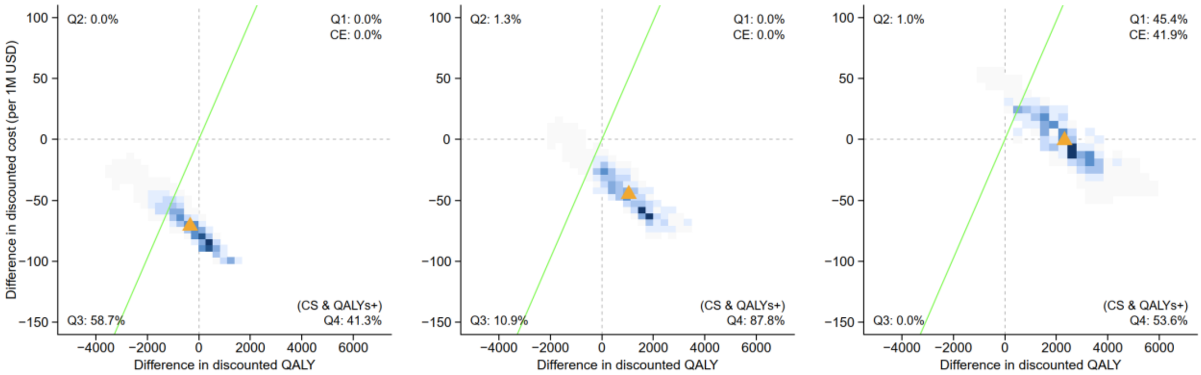

Abbreviations: CE, cost-effective; CS: cost-saving; QALY, quality-adjusted life year; Q1 to Q4, quadrant 1 to quadrant 4 of the cost-effectiveness plane; 1F1M, one-dose schedule for both schoolgirls and schoolboys; 2dFOV, two-dose schedule for female-only vaccination.

Notes. [(a)] The two-dose and one-dose vaccination schedules were assumed to provide lifelong and 20-year protection to vaccinees, respectively. [(b)] Vaccine uptake among girls was 85% in all GNV and FOV schedules. [(c)] The green lines present the willingness to pay (WTP) threshold at US$48,757/QALY. The orange triangles denote the mean of the differences in discounted cost and QALY. [(d)] On the density heatmaps (2-D histograms), grey/lighter blue represents fewer simulations and darker blue represents more simulations. [(6)] In each subplot, the percentage of simulations in each quadrant (i.e., Q1-Q4) is presented. Quadrant Q1 (North-east, more costs and more QALYs) includes the percentage of simulations that were cost-effective at the WTP threshold. Quadrant Q3 (South-west, fewer costs and fewer QALYs) includes the percentage of simulations that were considered adoptable if the willingness to accept a QALY lost equals the threshold of WTP for a QALY gained. All percentages are based on the total number of simulations.

**eFigure 5**. Cost-effectiveness planes for 1F1M vs 2dFOV by vaccine uptake among boys (columns) in the base case and alternative scenarios (rows) if the one-dose schedule provided 30 years of protection.

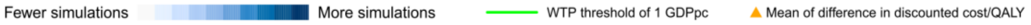

(A) Base case

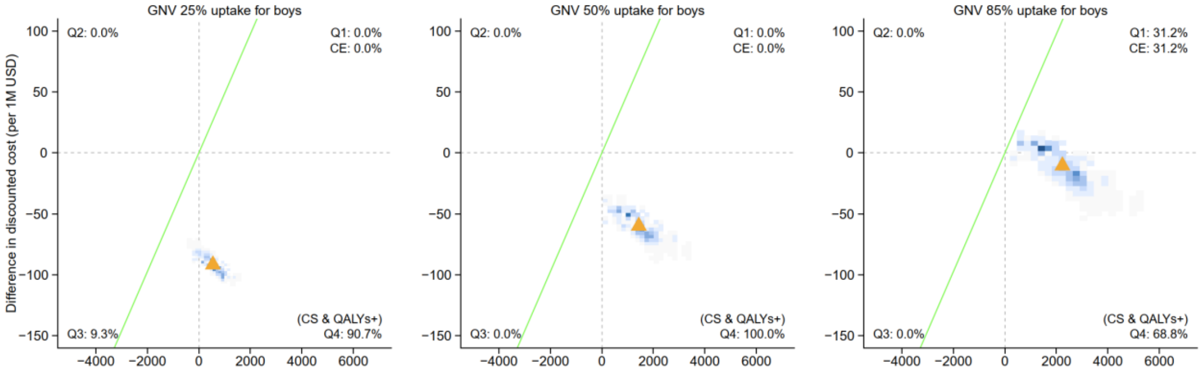

(B) 60% of HPV-related anal cancers in males were averted by FOV

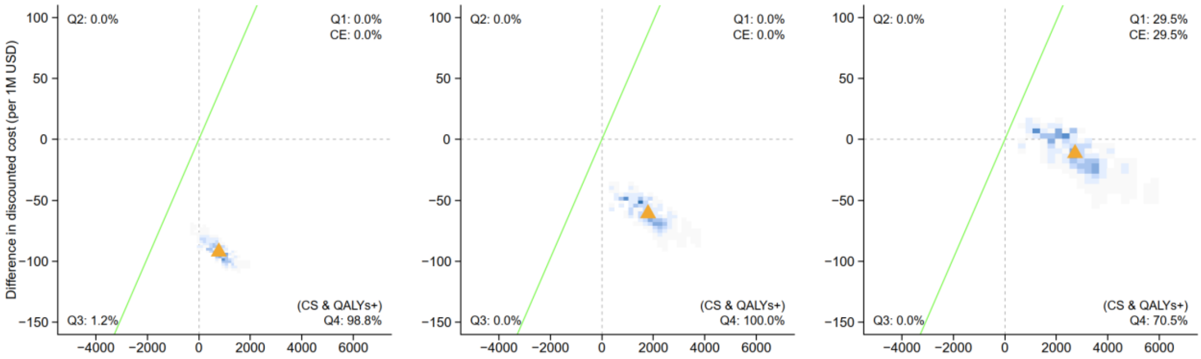

(C) Accounted for costs and QALYs associated with genital warts

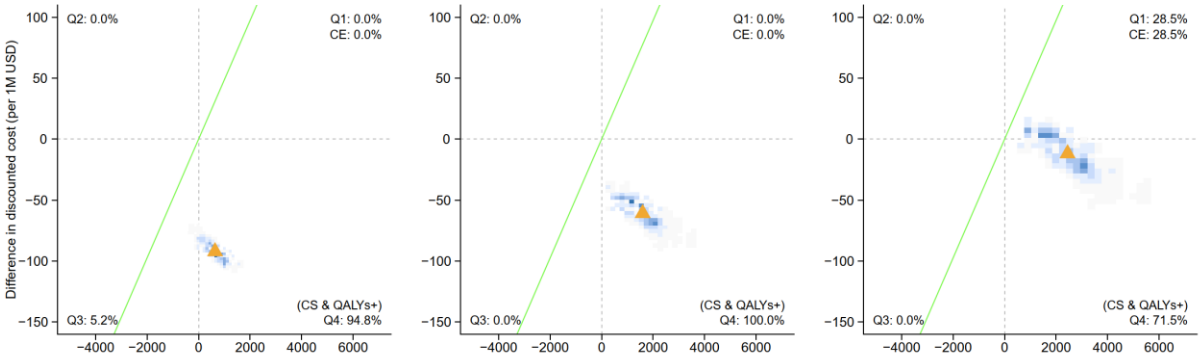

(D) Accounted for costs and QALYs associated with genital warts and 60% of HPV-related anal cancers in males were averted by FOV

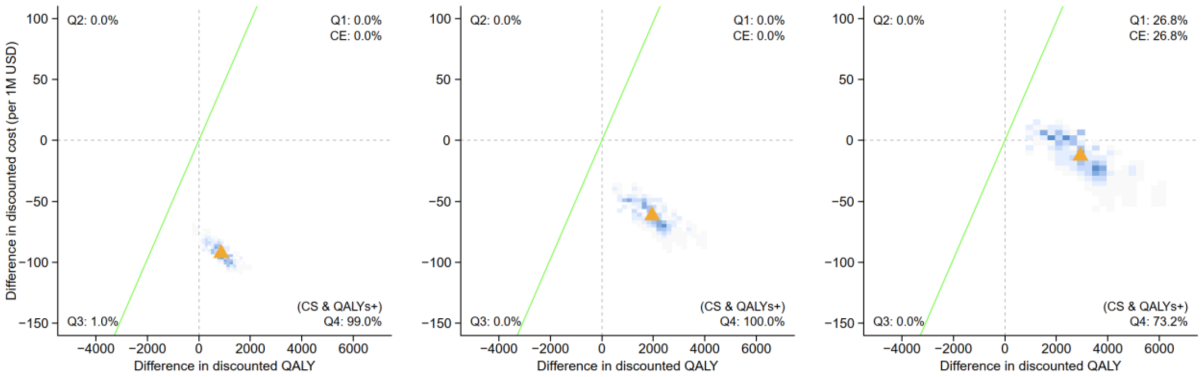

Abbreviations: CE, cost-effective; CS: cost-saving; QALY, quality-adjusted life year; Q1 to Q4, quadrant 1 to quadrant 4 of the cost-effectiveness plane; 1F1M, one-dose schedule for both schoolgirls and schoolboys; 2dFOV, two-dose schedule for female-only vaccination.

Notes. [(a)] The two-dose and one-dose vaccination schedules were assumed to provide lifelong and 30-year protection to vaccinees, respectively. [(b)] Vaccine uptake among girls was 85% in all GNV and FOV schedules. [(c)] The green lines present the willingness to pay (WTP) threshold at US$48,757/QALY. The orange triangles denote the mean of the differences in discounted cost and QALY. [(d)] On the density heatmaps (2-D histograms), grey/lighter blue represents fewer simulations and darker blue represents more simulations. [(e)] In each subplot, the percentage of simulations in each quadrant (i.e., Q1-Q4) is presented. Quadrant Q1 (North-east, more costs and more QALYs) includes the percentage of simulations that were cost-effective at the WTP threshold. Quadrant Q3 (South-west, fewer costs and fewer QALYs) includes the percentage of simulations that were considered adoptable if the willingness to accept a QALY lost equals the threshold of WTP for a QALY gained. All percentages are based on the total number of simulations.

**eFigure 6**. Incremental cost-effectiveness ratios (ICERs A) and threshold vaccination costs (TVCs B) for 2F1M vs 2dFOV by vaccine uptake among boys in the base case and alternative scenarios if the one-dose schedule provided 30-year protection to vaccinees.

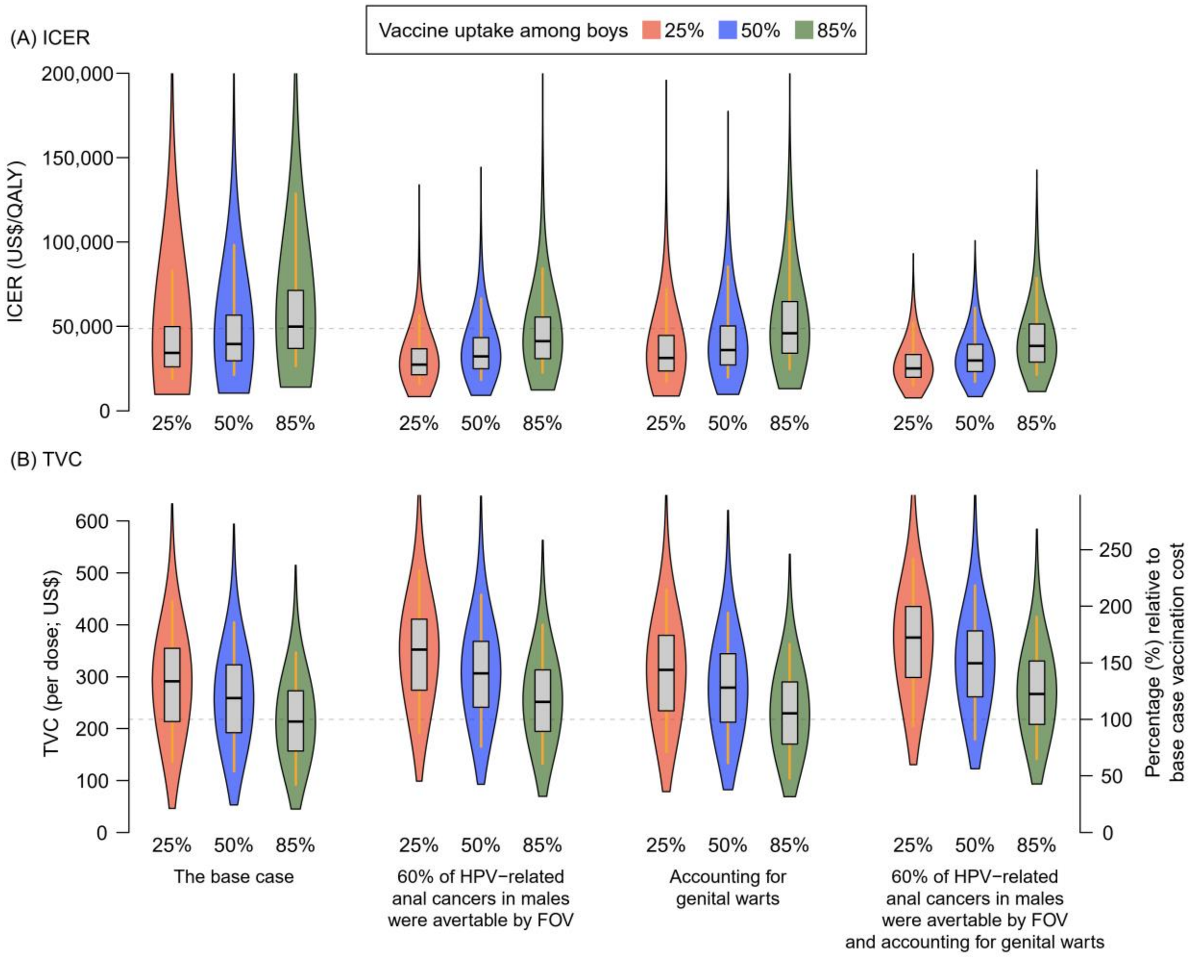

Abbreviations: FOV, female-only vaccination; GNV, gender-neutral vaccination; 2dFOV, FOV with a two-dose schedule for schoolgirls; 2F1M, two-dose vaccination schedule for schoolgirls and one-dose for schoolboys. Notes. (a) The two-dose and one-dose vaccination schedules were assumed to provide lifelong and 30-year protection to vaccinees, respectively. (b) Vaccine uptake among girls was 85% in all GNV and FOV strategies. (c) For panel (A), the willingness to pay (WTP) threshold was US$48,757 per QALY gain. For panel (B), the base case vaccination cost (vaccine cost plus administration expenses) was US$218 per dose. The WTP threshold and base case vaccination cost were presented by grey dashed lines on respective panels. (d) The violin plots present the smoothed kernel density of the values of ICERs/TVCs. The box plots present the medians (the horizontal lines), the 25th/75th percentiles (the boxes), and the 5th/95th percentiles (the whiskers).

**eFigure 7**. Incremental cost-effectiveness ratios (ICERs; A) and threshold vaccination costs (TVCs; B) for 2F1M vs 1F1M by vaccine uptake among boys in the base case and the scenario of accounting for genital warts if the 1one-dose schedule provided 20-year protection to vaccinees.

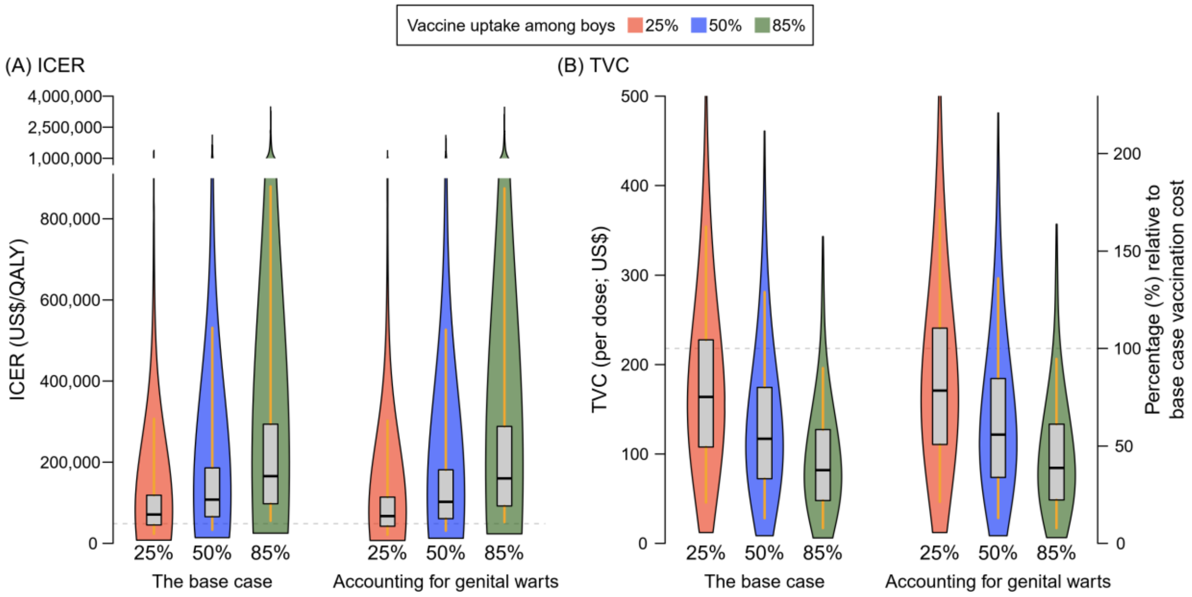

Abbreviations: FOV, female-only vaccination; GNV, gender-neutral vaccination; 1F1M, one-dose schedule for both schoolgirls and schoolboys; 2F1M, two-dose vaccination schedule for schoolgirls and one-dose for schoolboys.

Notes. [(a)] The two-dose and one-dose schedules were assumed to provide lifelong and 20-year protection to vaccinees, respectively. [(b)] Vaccine uptake among girls was 85% in all GNV and FOV strategies. [(c)] For panel (A), the willingness to pay (WTP) threshold was US$48,757 per QALY gain. For panel (B), the base case vaccination cost (vaccine cost plus administration expenses) was US$218 per dose. The WTP threshold and base case vaccination cost were presented by grey dashed lines on respective panels. [(d)] The violin plots present the smoothed kernel density of the values of ICERs/TVCs. The box plots present the medians (the horizontal lines), the 25$^{th}$/75$^{th}$ percentiles (the boxes), and the 5$^{th}$/95$^{th}$ percentiles (the whiskers). [(e)] Recalling that ICER is defined as the additional cost divided by the additional health benefit between two strategies. A high value of ICERs was observed if the difference in QALY gained between the two strategies is small. An axis-break is applied to display the extreme values of ICERs.

**eFigure 8**. Incremental cost-effectiveness ratios (ICERs; A) and threshold vaccination costs (TVCs; B) for 2F1M vs 1F1M by vaccine uptake among boys in the base case and the scenario of accounting for genital warts if the one-dose schedule provided 30-year protection to vaccinees.

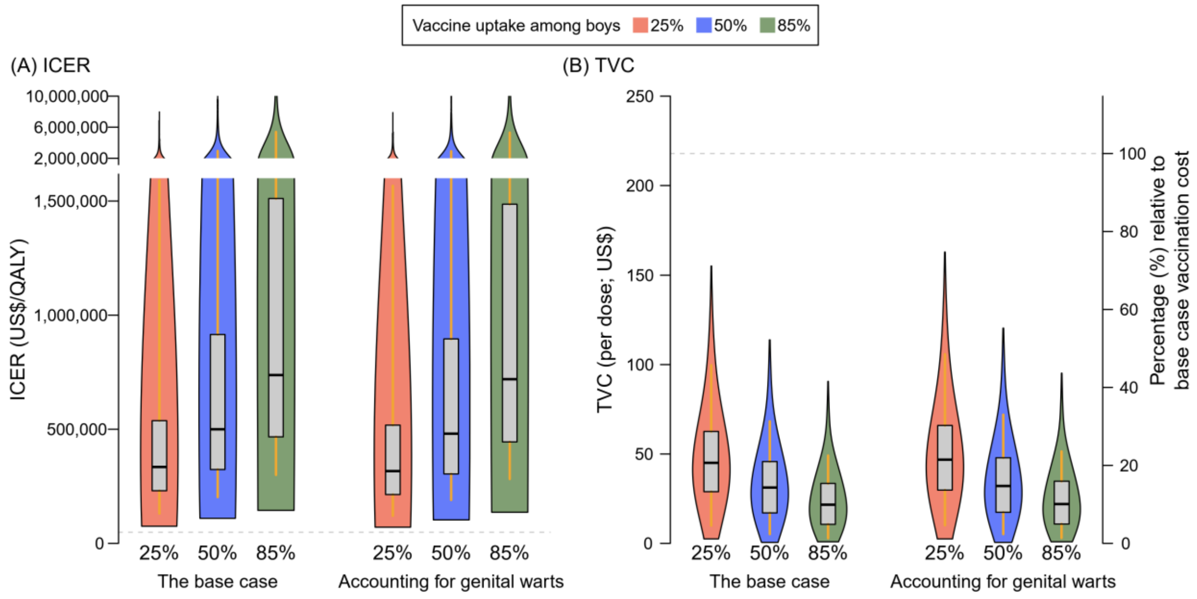

Abbreviations: FOV, female-only vaccination; GNV, gender-neutral vaccination; 2F1M, two-dose vaccination schedule for schoolgirls and one-dose for schoolboys; 1F1M, one-dose schedule for both schoolgirls and schoolboys.

Notes. [a] The two-dose and one-dose schedules were assumed to provide lifelong and 30-year protection to vaccinees, respectively. [b] Vaccine uptake among girls was 85% in all GNV and FOV strategies. [c] For panel (A), the willingness to pay (WTP) threshold was US$48,757 per QALY gain. For panel (B), the base case vaccination cost (vaccine cost plus administration expenses) was US$218 per dose. The WTP threshold and base case vaccination cost were presented by grey dashed lines on respective panels. [d] The violin plots present the smoothed kernel density of the values of ICERs/TVCs. The box plots present the medians (the horizontal lines), the 25$^{th}$/75$^{th}$ percentiles (the boxes), and the 5$^{th}$/95$^{th}$ percentiles (the whiskers). [e] Recalling that ICER is defined as the additional cost divided by the additional health benefit between two strategies. A high value of ICERs is observed if the difference in QALY gained between the two strategies is small. An axis-break is applied to display the extreme values of ICERs.

**eFigure 9**. Distribution of the difference in cost (A) and QALY gained (B) that were associated with treatment to health outcomes of HPV-related cancers and genital warts for 2F2M vs 2dFOV.

(A)(i) [The base case] Difference in cost for 2F2M vs 2dFOV

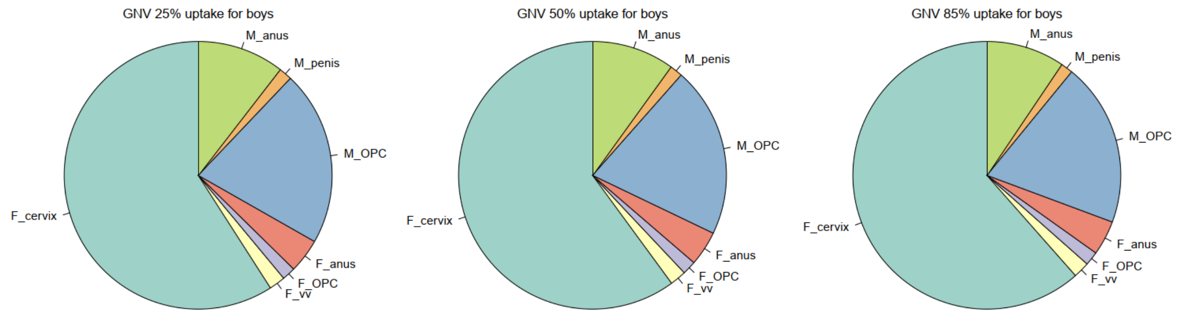

(A)(ii) [The base case] Difference in QALY gained for 2F2M vs 2dFOV

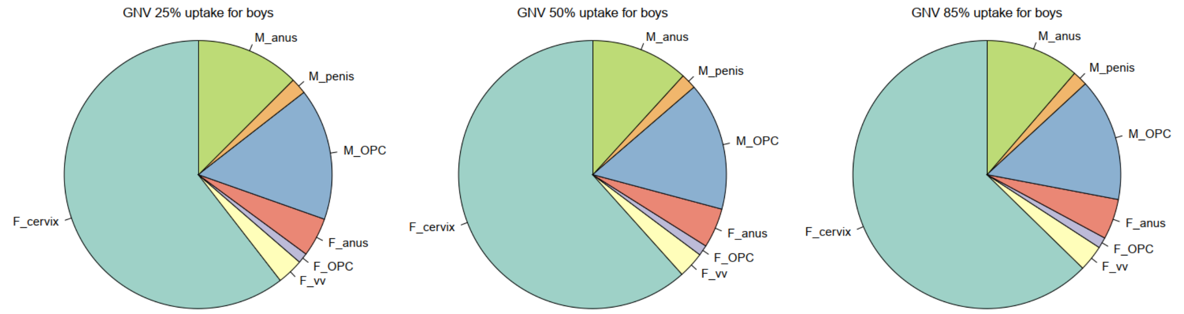

(B)(i) [Accounting for genital warts] Difference in cost for 2F2M vs 2dFOV

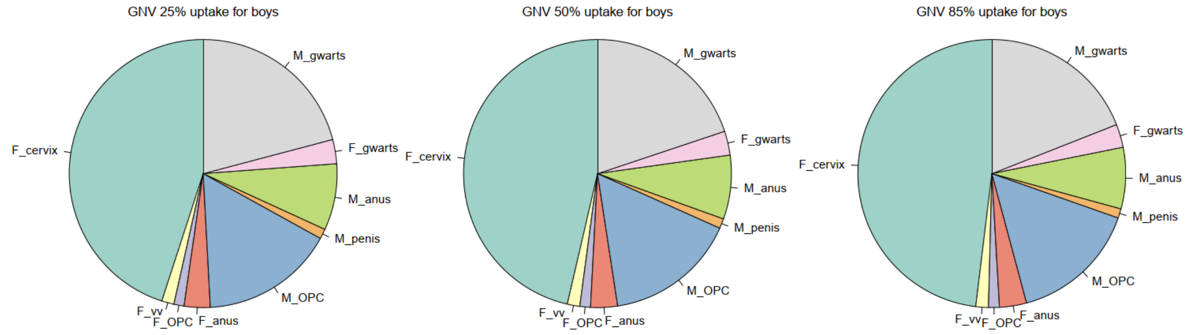

(B)(ii) [Accounting for genital warts] Difference in QALY gained for 2F2M vs 2dFOV

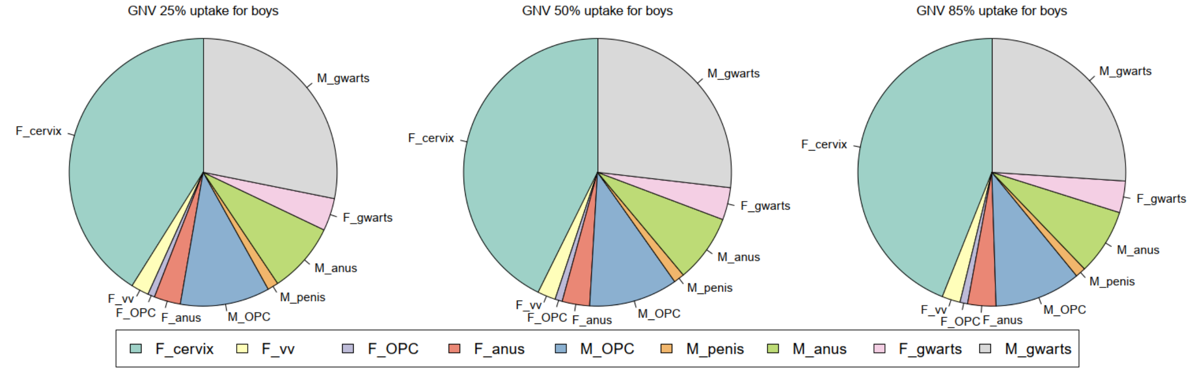

Abbreviations: F_dis and M_dis, the disease related to females and males, respectively; gwarts, genital warts; OPC, oropharyngeal cancer; vv, vaginal/vulvar cancer; 2dFOV, two-dose schedule for female-only vaccination; 2F2M, gender-neutral vaccination with two-dose schedule for schoolgirls and schoolboys.

Notes. The pie charts present the distribution of the median of the difference in cost and QALY gained associated with the treatment of health outcomes of HPV-related cancers and/or genital warts. Assumed that the two-dose schedule provided lifelong protection.

**eFigure 10**. Distribution of the difference in cost (A) and QALY gained (B) associated with treatment to health outcomes of HPV-related cancers and genital warts for 2F2M vs 2dFOV.

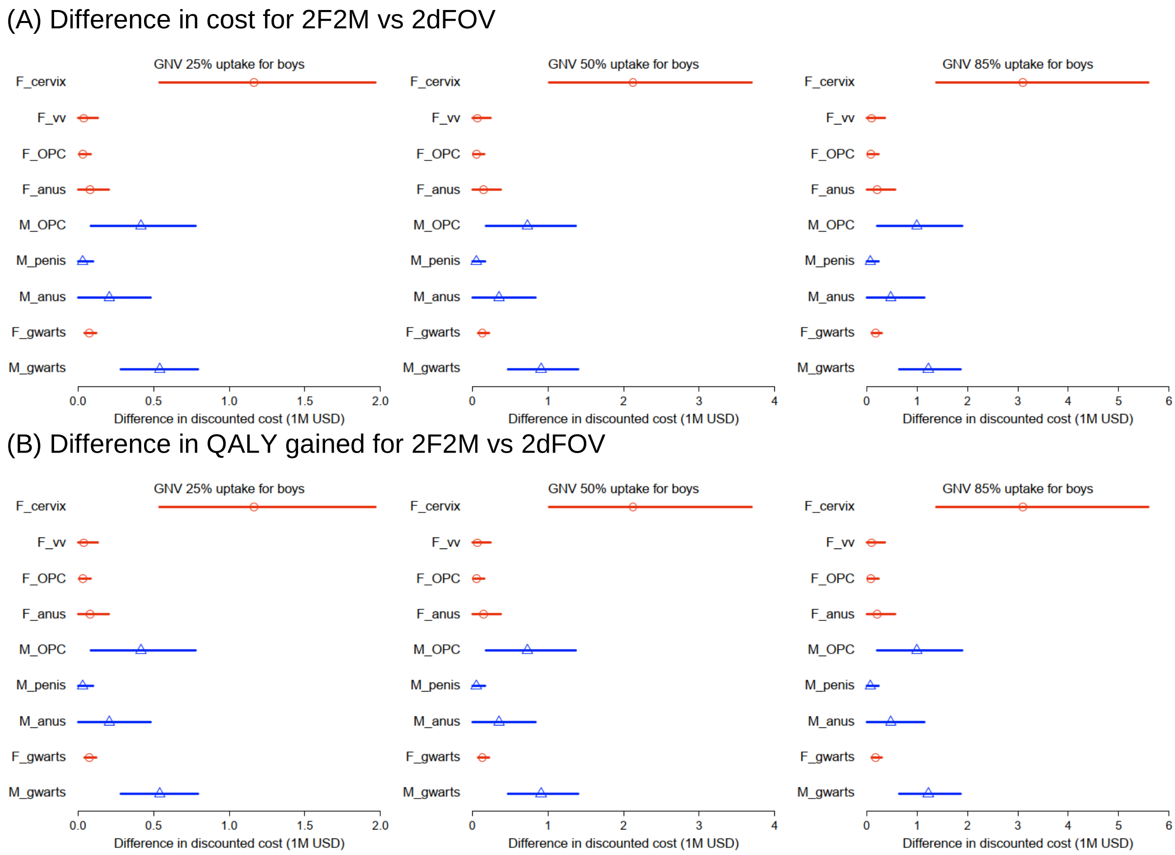

Abbreviations: F_dis and M_dis, the disease related to females and males, respectively; gwarts, genital warts; OPC, oropharyngeal cancer; vv, vaginal/vulvar cancer; 2dFOV, two-dose schedule for female-only vaccination; 2F2M, two-dose vaccination schedule for schoolgirls and schoolboys.
Notes. The markers and the lines present the medians and the 90% prediction intervals ($5^{th}$ to $95^{th}$ percentiles), respectively, of the difference in cost and QALY gained associated with the treatment of health outcomes of HPV-related cancers and/or genital warts. Assumed that the two-dose schedule provided lifelong protection.

**eFigure 11**. Distribution of the difference in cost (A) and QALY gained (B) associated with treatment to health outcomes of HPV-related cancers and genital warts for 1F1M vs 2dFOV if one-dose schedule was assumed to provide 20-year protection.

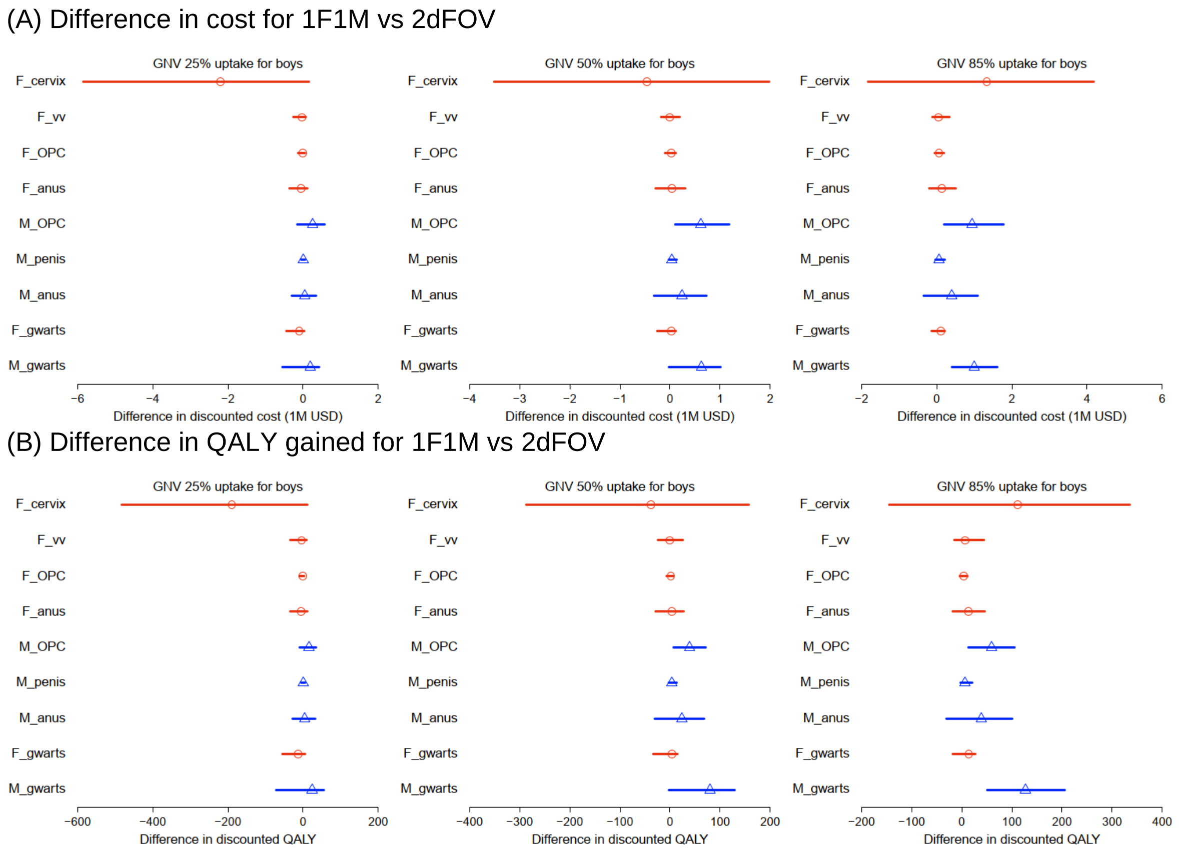

Abbreviations: F_dis and M_dis, the disease related to females and males, respectively; gwarts, genital warts; OPC, oropharyngeal cancer; vv, vaginal/vulvar cancer; 1F1M, one-dose vaccination schedule for schoolgirls and schoolboys; 2dFOV, two-dose schedule for femaly-only vaccination.

Notes. The markers and the lines present the medians and the 90% prediction intervals ($5^{th}$ to $95^{th}$ percentiles), respectively, of the difference in cost and QALY gained associated with the treatment of health outcomes of HPV-related cancers and/or genital warts.

**eFigure 12**. Distribution of the difference in cost (A) and QALY gained (B) associated with treatment to health outcomes of HPV-related cancers and genital warts for 1F1M vs 2dFOV if one-dose schedule was assumed to provide 30-year protection.

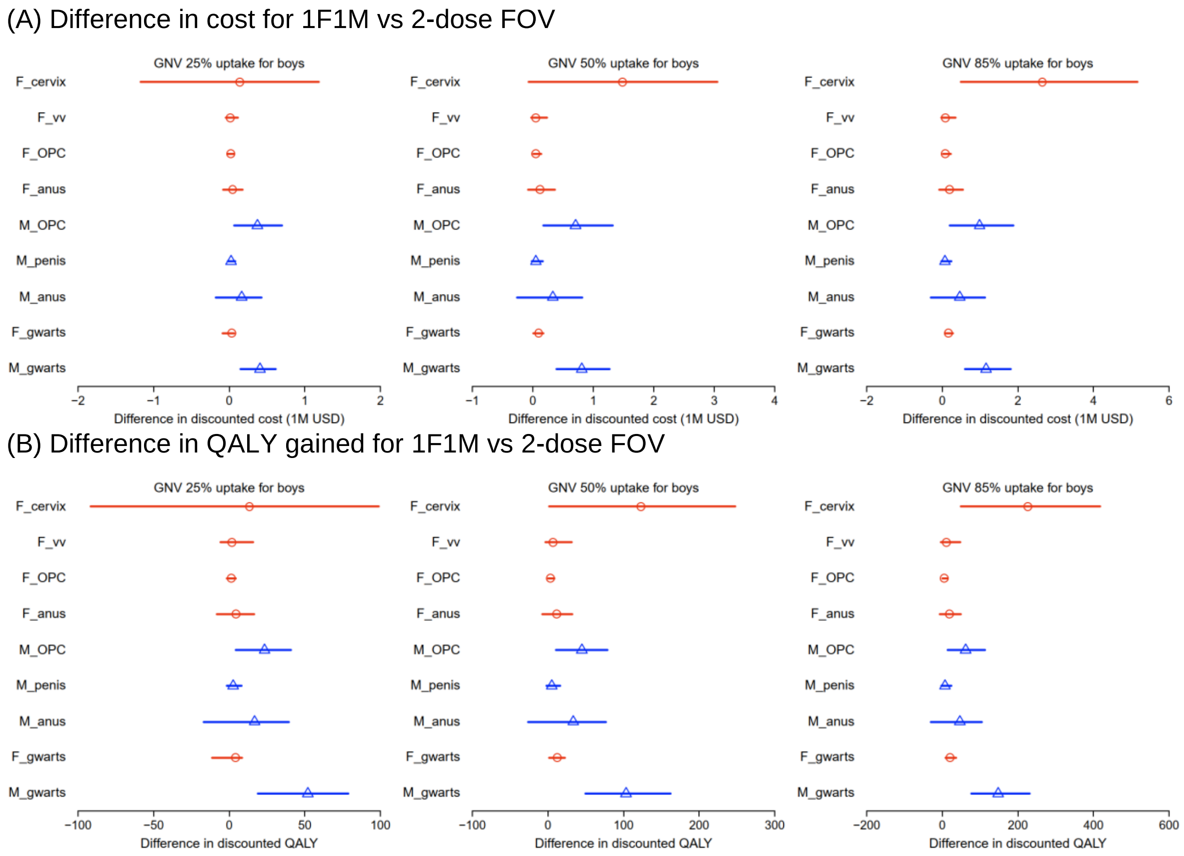

Abbreviations: F_dis and M_dis, the disease related to females and males, respectively; gwarts, genital warts; OPC, oropharyngeal cancer; vv, vaginal/vulvar cancer; 1F1M, one-dose vaccination schedule for schoolgirls and schoolboys; 2dFOV, two-dose schedule for female-only vaccination.
Notes. The markers and the lines present the medians and the 90% prediction intervals ($5^{th}$ to $95^{th}$ percentiles), respectively, of the difference in cost and QALY gained associated with the treatment of health outcomes of HPV-related cancers and/or genital warts.

**eFigure 13**. HPV-16 infection among females by vaccine uptake in schoolboys and protection duration of the one-dose schedule.

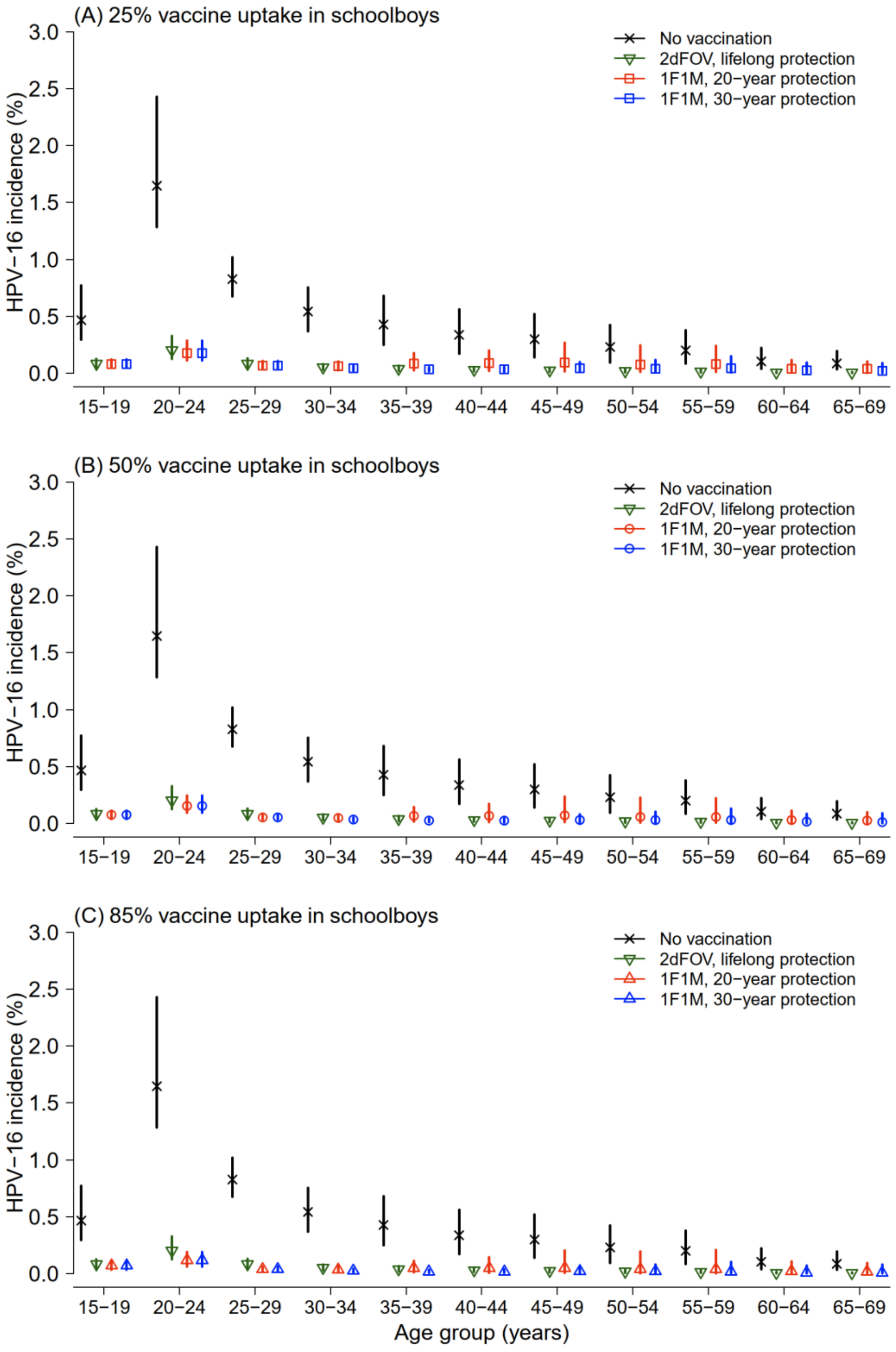

Abbreviations: 1F1M, one-dose schedule for both genders; 2dFOV, two-dose schedule for female-only vaccination.

Notes. [(a)] The markers and the lines present the medians and the 90% prediction interval ($5^{th}$ to $95^{th}$ percentiles). [(b)] Vaccine uptake among schoolgirls was assumed to be 85% for 2dFOV and 1F1M. [(c)] HPV incidence in 2dFOV and 1F1M was based on the incidence in the first vaccinated cohort of implementing GNV (age 12 years old in 2024).

**eFigure 14.** HPV-16 infection among males by vaccine uptake in schoolboys and protection duration of the one-dose schedule.

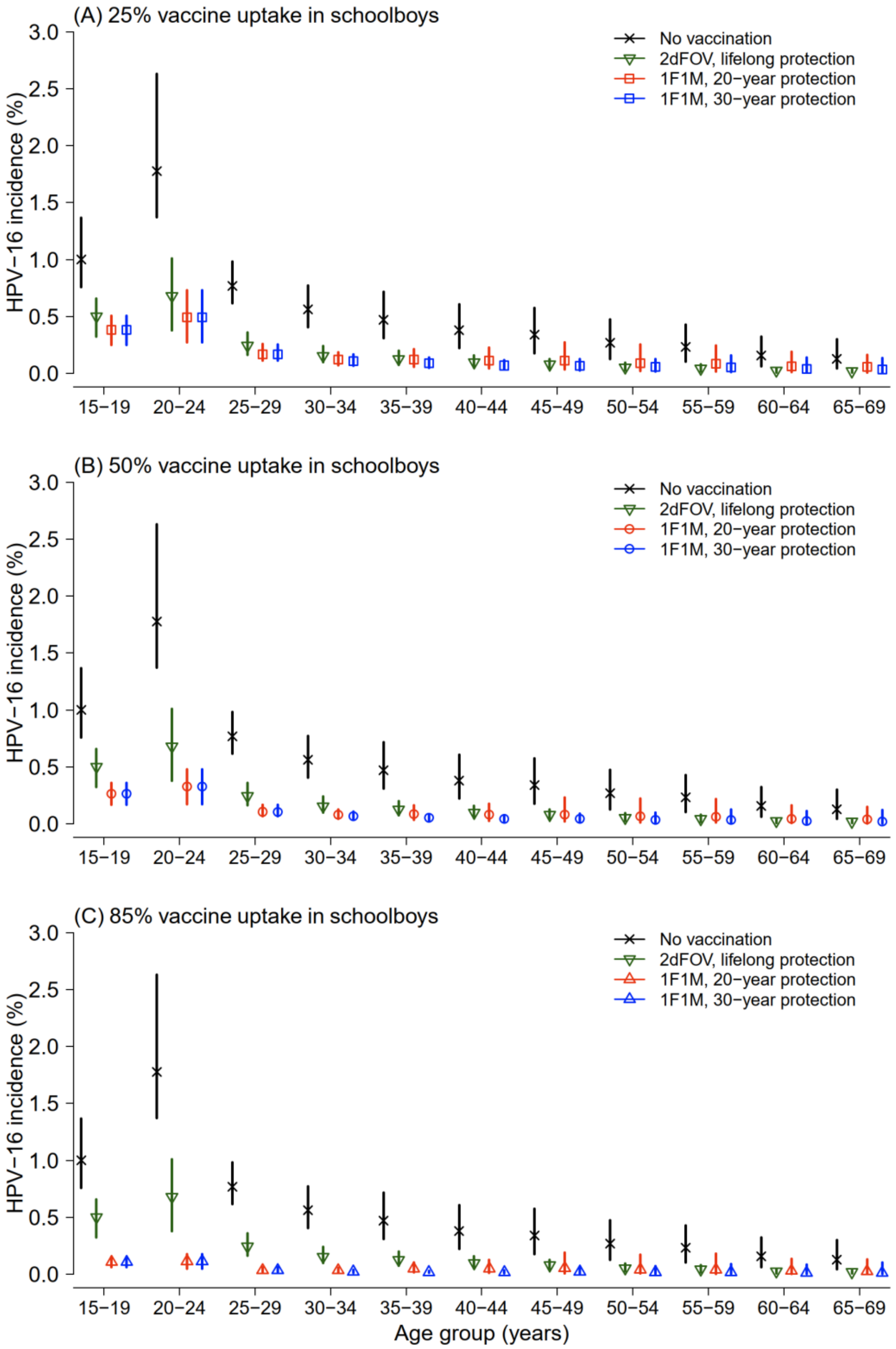

Abbreviations: 1F1M, one-dose schedule for both genders; 2dFOV, two-dose schedule for female-only vaccination.

Notes. (a) The markers and the lines present the medians and the 90% prediction interval (5th to 95th percentiles). (b) Vaccine uptake among schoolgirls was assumed to be 85% for 2dFOV and 1F1M. (c) HPV incidence in 2dFOV and 1F1M was based on the incidence in the first vaccinated cohort of implementing GNV (age 12 years old in 2024).

**eTable 3**. ICERs of 2F2M vs 2dFOV and the respective TVCs by vaccine uptake among boys in the base case and alternative scenarios. [a]

| Vaccine uptake among boys [b] | ICER (US$/QALY) [Median (90% PI)] | TVC (US$) [Median (90% PI)] | Percentage of simulations in which ICERs were below the WTP threshold at the base case vaccination cost [c,d] | Relative reduction (%) of vaccination cost to its base case so that 80%/85%/90%/95% of simulations were cost-effective at the WTP threshold [c,d] |
|---|---|---|---|---|
| (A) Base case | | | | |
| 25% | 77,926 (47,368, 175,267) | 145.8 (68.5, 223.4) | 5.5% | 54.4 / 58.0 / 62.7 / 68.6 |
| 50% | 88,892 (53,106, 204,564) | 129.5 (59.0, 202.8) | 3.4% | 58.7 / 61.6 / 65.9 / 72.9 |
| 85% | 109,375 (63,824, 264,980) | 106.9 (46.4, 173.6) | 0.9% | 66.1 / 68.7 / 71.9 / 78.7 |
| (B) 60% of HPV-related anal cancers in males were avertable by FOV | | | | |
| 25% | 61,940 (40,908, 117,919) | 176.5 (96.8, 253.4) | 15.6% | 41.0 / 44.6 / 48.8 / 55.6 |
| 50% | 72,453 (45,911, 138,012) | 153.5 (83.0, 229.3) | 5.6% | 47.8 / 51.5 / 55.8 / 61.9 |
| 85% | 89,618 (53,987, 174,436) | 125.9 (66.3, 200.1) | 1.8% | 57.4 / 60.2 / 64.7 / 69.6 |
| (C) Accounted for costs and QALYs associated with genital warts | | | | |
| 25% | 71,145 (44,398, 151,053) | 157.6 (77.9, 235.0) | 15.6% | 49.2 / 53.4 / 58.7 / 64.3 |
| 50% | 81,975 (50,371, 179,057) | 139.6 (67.0, 212.1) | 5.6% | 55.0 / 57.9 / 62.6 / 69.2 |
| 85% | 101,243 (60,735, 232,608) | 114.6 (52.5, 181.7) | 1.8% | 63.1 / 65.7 / 69.3 / 75.9 |
| (D) 60% of HPV-related anal cancers in males were avertable by FOV and accounted for costs and QALYs associated with genital warts | | | | |
| 25% | 57,474 (38,816, 109,264) | 188.6 (103.6, 264.5) | 26.3% | 36.2 / 39.6 / 44.8 / 52.5 |
| 50% | 67,797 (43,670, 128,110) | 163.1 (90.2, 238.3) | 8.4% | 43.8 / 47.3 / 52.2 / 58.6 |
| 85% | 84,247 (51,560, 163,415) | 133.6 (71.2, 207.9) | 3.3% | 53.7 / 57.2 / 61.7 / 67.3 |

Abbreviations: ICER, incremental cost-effectiveness ratio; PI, prediction interval (5$^{th}$ to 95$^{th}$ percentiles for 90% PI); QALY, quality-adjusted life year; TVC, threshold vaccination cost (vaccine cost plus administration expenses) for each dose; WTP, willingness to pay; 2dFOV, two-dose schedule for female-only vaccination; 2F2M, two-dose schedule for both schoolgirls and schoolboys.

Notes. [(a)] We assumed that the two-dose schedule provided lifelong vaccine-induced protection. [(b)] Vaccine uptake among girls was 85% in all GNV and FOV schedules. [(c)] The WTP threshold was set at US$48,757 per QALY gain. [(d)] The base case vaccination cost (vaccine cost plus administration expenses) was US$218 per dose.

**eTable 4**. ICERs of 2F1M vs 2dFOV and the respective TVCs by vaccine uptake among boys in the base case and alternative scenarios if the one-dose schedule provided 20-year protection. [a]

| Vaccine uptake among boys [b] | ICER (US$/QALY) [Median (90% PI)] | TVC (US$) [Median (90% PI)] | Percentage of simulations in which ICERs were below the WTP threshold at the base case vaccination cost [c,d] | Relative reduction (%) of vaccination cost to its base case so that 80%/85%/90%/95% of simulations were cost-effective at the WTP threshold [c,d,e] |
|---|---|---|---|---|
| (A) Base case | | | | |
| 25% | 34,831 (18,907, 85,115) | 286.8 (133.4, 442.1) | 73.1% | 12.2 / 17.2 / 26.4 / 38.8 |
| 50% | 39,816 (21,449, 100,056) | 257.3 (116.5, 403.6) | 63.4% | 18.6 / 24.0 / 32.0 / 46.6 |
| 85% | 50,285 (26,541, 129,534) | 212.4 (91.7, 345.9) | 47.2% | 33.3 / 37.9 / 44.0 / 57.9 |
| (B) 60% of HPV-related anal cancers in males were avertable by FOV | | | | |
| 25% | 27,724 (15,854, 57,923) | 348.7 (186.4, 501.9) | 90.8% | −14.7 / −8.7 / −1.3 / 14.5 |
| 50% | 32,582 (18,328, 67,506) | 305.0 (162.9, 453.2) | 82.0% | −3.3 / 3.8 / 12.8 / 25.3 |
| 85% | 41,463 (22,677, 84,928) | 250.2 (131.3, 397.6) | 64.4% | 15.7 / 21.1 / 29.5 / 39.7 |
| (C) Accounted for costs and QALYs associated with genital warts | | | | |
| 25% | 31,678 (17,404, 73,816) | 310.4 (151.2, 467.1) | 79.1% | 1.6 / 9.0 / 18.6 / 30.6 |
| 50% | 36,311 (20,025, 87,036) | 276.9 (131.4, 422.5) | 72.4% | 11.7 / 16.9 / 25.8 / 39.7 |
| 85% | 46,353 (24,775, 112,994) | 227.6 (103.6, 361.9) | 53.9% | 27.4 / 32.1 / 39.0 / 52.4 |
| (D) 60% of HPV-related anal cancers in males were avertable by FOV and accounted for costs and QALYs associated with genital warts | | | | |
| 25% | 25,527 (14,718, 53,521) | 371.7 (200.6, 522.8) | 93.7% | −24.2 / −17.9 / −9.0 / 7.9 |
| 50% | 30,068 (17,160, 61,628) | 324.2 (177.5, 474.2) | 87.9% | −11.0 / −3.8 / 5.3 / 18.6 |
| 85% | 38,674 (21,449, 79,306) | 265.3 (140.6, 414.5) | 70.0% | 8.2 / 15.4 / 23.4 / 35.5 |

Abbreviations: ICER, incremental cost-effectiveness ratio; PI, prediction interval (5th to 95th percentiles for 90% PI); QALY, quality-adjusted life year; TVC, threshold vaccination cost (vaccine cost plus administration expenses) for each dose; WTP, willingness to pay; 2dFOV, two-dose schedule for female-only vaccination; 2F1M, two-dose schedule for schoolgirls and one-dose for schoolboys.

Notes. (a) The two-dose schedule for schoolgirls and one-dose schedule for schoolboys were assumed to provide lifelong and 20-year vaccine-induced protection, respectively. (b) Vaccine uptake among girls was 85% in all vaccination schedules. (c) The WTP threshold was set at US$48,757 per QALY gain. (d) The base case vaccination cost (vaccine cost plus administration expenses) was US$218 per dose. (e) A negative relative reduction of vaccination cost refers to the relative increase in the base case vaccination cost when 2F1M was cost-effective with an ICER below the WTP threshold. In this case, a higher vaccination cost would be acceptable for the intervention and the comparator to be equally cost-effective at the WTP threshold.

**eTable 5**. ICERs of 2F1M vs 2dFOV and the respective TVCs by vaccine uptake among boys in the base case and alternative scenarios if the one-dose schedule provided 30-year protection. [a]

| Vaccine uptake among boys [b] | ICER (US$/QALY) [Median (90% PI)] | TVC (US$) [Median (90% PI)] | Percentage of simulations in which ICERs were below the WTP threshold at the base case vaccination cost [c,d] | Relative reduction (%) of vaccination cost to its base case so that 80%/85%/90%/95% of simulations were cost-effective at the WTP threshold [c,d,e] |
|---|---|---|---|---|
| (A) Base case | | | | |
| 25% | 34,323 (18,631, 83,109) | 291.1 (136.3, 445.7) | 73.4% | 9.7 /16.5 / 25.3 / 37.5 |
| 50% | 39,535 (21,398, 98,211) | 258.8 (117.8, 405.0) | 65.0% | 17.3 / 23.4 / 32.2 / 46.0 |
| 85% | 49,876 (26,412, 128,624) | 213.8 (91.9, 346.8) | 47.9% | 32.4 / 37.4 / 43.8 / 57.8 |
| (B) 60% of HPV-related anal cancers in males were avertable by FOV | | | | |
| 25% | 27,410 (15,840, 56,343) | 352.2 (191.6, 505.1) | 91.1% | −17.1 / −9.8 / −2.8 / 12.1 |
| 50% | 32,284 (18,325, 66,400) | 306.8 (165.2, 458.1) | 82.7% | −4.4 / 3.1 / 12.5 / 24.2 |
| 85% | 41,197 (22,604, 84,261) | 251.6 (132.2, 399.8) | 65.3% | 15.0 / 20.4 / 29.2 / 39.3 |
| (C) Accounted for costs and QALYs associated with genital warts | | | | |
| 25% | 31,267 (17,159, 72,004) | 313.4 (154.3, 468.1) | 80.7% | −1.2 / 7.9 / 16.9 / 29.2 |
| 50% | 36,053 (19,853, 85,400) | 279.0 (133.4, 423.3) | 73.4% | 10.2 / 16.0 / 25.9 / 38.8 |
| 85% | 45,899 (24,642, 112,140) | 229.4 (104.5, 362.8) | 54.4% | 26.5 / 31.6 / 38.7 / 52.0 |
| (D) 60% of HPV-related anal cancers in males were avertable by FOV and accounted for costs and QALYs associated with genital warts | | | | |
| 25% | 25,145 (14,704, 52,159) | 375.5 (205.1, 527.2) | 93.9% | −26.7 / −19.7 / −10.6 / 5.9 |
| 50% | 29,731 (17,141, 60,940) | 326.1 (179.3, 475.6) | 88.1% | −12.2 / −5.2 / 4.9 / 17.7 |
| 85% | 38,380 (21,357, 78,590) | 267.1 (141.6, 415.4) | 70.6% | 7.7 / 14.4 / 23.2 / 35.0 |

Abbreviations: ICER, incremental cost-effectiveness ratio; PI, prediction interval ($5^{th}$ to $95^{th}$ percentiles for 90% PI); QALY, quality-adjusted life year; TVC, threshold vaccination cost (vaccine cost plus administration expenses) for each dose; WTP, willingness to pay; 2dFOV, two-dose schedule for female-only vaccination; 2F1M, two-dose schedule for schoolgirls and one-dose for schoolboys.

Notes. (a) The two-dose schedule for schoolgirls and one-dose schedule for schoolboys were assumed to provide lifelong and 30-year vaccine-induced protection, respectively. (b) Vaccine uptake among girls was 85% in all vaccination schedules. (c) The WTP threshold was set at US$48,757 per QALY gain. (d) The base case vaccination cost (vaccine cost plus administration expenses) was US$218 per dose. (e) A negative relative reduction of vaccination cost refers to the relative increase compared to the base case vaccination cost when 2F1M was cost-effective with an ICER below the WTP threshold. In this case, a higher vaccination cost would be acceptable for the intervention and the comparator to be equally cost-effective at the WTP threshold.

**eTable 6**. ICERs of 2F1M vs 1F1M and the respective TVCs by vaccine uptake among boys in the base case and the scenario of accounting for genital warts if the one-dose schedule provided 20-year protection. [a]

| Vaccine uptake among boys [b] | ICER (US$/QALY) [Median (90% PI)] | TVC (US$) [Median (90% PI)] | Percentage of simulations in which ICERs were below the WTP threshold at the base case vaccination cost [c,d] | Relative reduction (%) of vaccination cost to its base case so that 80%/85%/90%/95% of simulations were cost-effective at the WTP threshold |
|---|---|---|---|---|
| (A) Base case | | | | |
| 25% | 71,144 (23,453, 307,123) | 164.0 (45.9, 354.2) | 28.4% | 54.5 / 59.8 / 67.6 / 78.9 |
| 50% | 108,191 (34,214, 531,219) | 117.2 (28.2, 281.1) | 13.3% | 70.3 / 72.7 / 76.9 / 87.0 |
| 85% | 166,080 (56,051, 880,060) | 82.0 (17.2, 196.2) | 3.3% | 80.4 / 83.0 / 85.6 / 92.1 |
| (B) Accounted for costs and QALYs associated with genital warts | | | | |
| 25% | 67,364 (21,601, 302,276) | 170.9 (46.6, 372.8) | 31.6% | 53.4 / 58.8 / 67.0 / 78.6 |
| 50% | 102,915 (31,691, 526,934) | 121.7 (28.5, 296.3) | 15.3% | 69.7 / 72.3 / 76.5 / 86.9 |
| 85% | 160,305 (52,325, 875,128) | 84.3 (17.3, 206.4) | 3.8% | 80.1 / 82.9 / 85.4 / 92.1 |

Abbreviations: ICER, incremental cost-effectiveness ratio; PI, prediction interval ($5^{th}$ to $95^{th}$ percentiles for 90% PI); QALY, quality-adjusted life year; TVC, threshold vaccination cost (vaccine cost plus administration expenses) for each dose; WTP, willingness to pay; 1F1M, one-dose schedule for both schoolgirls and schoolboys; 2F1M, two-dose schedule for schoolgirls and one-dose for schoolboys.

Notes. [(a)] The two-dose and one-dose schedules were assumed to provide lifelong and 20-year vaccine-induced protection, respectively. [(b)] Vaccine uptake among girls was 85% in all GNV and FOV schedules. [(c)] The WTP threshold was set at US$48,757 per QALY gain. [(d)] The base case vaccination cost (vaccine cost plus administration expenses) was US$218 per dose.

**eTable 7**. ICERs of 2F1M vs 1F1M and the respective TVCs by vaccine uptake among boys in the base case and the scenario of accounting for genital warts if the one-dose schedule provided 30-year protection. [a]

| Vaccine uptake among boys [b] | ICER (US$/QALY) [Median (90% PI)] | TVC (US$) [Median (90% PI)] | Percentage of simulations in which ICERs were below the WTP threshold at the base case vaccination cost [c,d] | Relative reduction (%) of vaccination cost to its base case so that 80%/85%/90%/95% of simulations were cost-effective at the WTP threshold [c,d] |
|---|---|---|---|---|
| (A) Base case | | | | |
| 25% | 335,732 (130,715, 1,590,018) | 45.0 (10.0, 99.7) | 0% | 88.2 / 90.2 / 91.7 / 95.4 |
| 50% | 501,039 (203,727, 2,939,704) | 31.2 (5.2, 68.2) | 0% | 93.0 / 93.7 / 94.7 / 97.6 |
| 85% | 738,518 (299,904, 5,370,149) | 21.7 (3.0, 49.2) | 0% | 95.6 / 96.1 / 96.9 / 98.6 |
| (B) Accounted for costs and QALYs associated with genital warts | | | | |
| 25% | 316,981 (120,770, 1,566,678) | 46.9 (10.1, 105.8) | 0% | 88.0 / 90.0 / 91.5 / 95.3 |
| 50% | 481,261 (190,420, 2,913,082) | 32.1 (5.2, 72.0) | 0% | 92.8 / 93.6 / 94.5 / 97.6 |
| 85% | 719,728 (282,651, 5,316,288) | 22.1 (3.0, 51.4) | 0% | 95.5 / 96.0 / 96.9 / 98.6 |

Abbreviations: ICER, incremental cost-effectiveness ratio; PI, prediction interval (5$^{th}$ to 95$^{th}$ percentiles for 90% PI); QALY, quality-adjusted life year; TVC, threshold vaccination cost (vaccine cost plus administration expenses) for each dose; WTP, willingness to pay; 1F1M, one-dose schedule for both schoolgirls and schoolboys; 2F1M, two-dose schedule for schoolgirls and one-dose for schoolboys.

Notes. (a) The two-dose and one-dose schedules were assumed to provide lifelong and 30-year vaccine-induced protection, respectively. (b) Vaccine uptake among girls was 85% in all GNV and FOV schedules. (c) The WTP threshold was set at US$48,757 per QALY gain. (d) The base case vaccination cost (vaccine cost plus administration expenses) was US$218 per dose.

**eTable 8**. The cost-effective vaccination strategy in two scenarios around vaccination cost in the situation that considers a GNV strategy effective if it incurred more QALYs than the status quo 2dFOV in at least 80% of simulations. [a]

| (A) At the base case vaccination cost | | | |
|---|---|---|---|
| | Vaccine uptake among boys [b] | | |
| Duration of vaccine-induced protection by the one-dose schedule | 25% | 50% | 85% |
| 20-year | 2dFOV [c] | 2dFOV | 1F1M |
| 30-year | 1F1M | 1F1M | 1F1M |
| (B) If the vaccination cost could be moderately reduced [d] | | | |
| | Vaccine uptake among boys | | |
| Duration of vaccine-induced protection by the one-dose schedule | 25% | 50% | 85% |
| 20-year | 2F1M (12%) | 2F1M (19%) | 1F1M |
| 30-year | 1F1M | 1F1M | 1F1M |

Abbreviations: FOV, female-only vaccination; GNV, gender-neutral vaccination; QALY, quality-adjusted life year; 1F1M, one-dose schedule for both schoolgirls and schoolboys; 2dFOV, two-dose schedule for FOV; 2F1M, two-dose schedule for schoolgirls and one-dose schedule for schoolboys.

Notes. (a) The findings were based on the base case scenario that assumed all HPV-related anal cancers in males are equally avertable by FOV via indirect protection and did not take into account of the impact of HPV vaccination on genital warts. (b) Vaccine uptake among schoolgirls was assumed to be 85% in all strategies. (c) The two-dose schedule was assumed to provide lifelong protection. (d) For panel (B), we considered the alternative strategies acceptable if the vaccination cost could be moderately reduced by no more than 50%. In this setting, the numbers in parentheses present the relative reduction in vaccination cost (vaccine cost plus administration expenses) for the alternative strategy to be cost-effective in 80% of simulations at the threshold of US$48,757 per QALY gain.

**eTable 9**. Additional relative reduction in the number of genital warts prevented by GNV strategies compared to that by the status quo 2dFOV. [a, b]

| GNV strategies | Vaccine uptake among boys in GNV strategies | Additional relative reduction in females | Additional relative reduction in males |
|---|---|---|---|
| (A) 2F2M [c] | 25% | 1.7% (0.5%, 2.9%) [d] | 4.8% (1.6%, 7.7%) |
| | 50% | 2.6% (0.8%, 5.6%) | 6.9% (2.5%, 13.2%) |
| | 85% | 3.2% (1.2%, 6.7%) | 8.4% (3.4%, 16.1%) |
| (B) Assuming that the one-dose schedule provided 20-year protection [c] | | | |
| (B)(i) 2F1M | 25% | 1.6% (0.5%, 2.8%) | 4.5% (1.5%, 7.3%) |
| | 50% | 2.5% (0.8%, 5.5%) | 6.8% (2.4%, 12.7%) |
| | 85% | 3.2% (1.2%, 6.6%) | 8.3% (3.1%, 15.9%) |
| (B)(ii) 1F1M | 25% | −1.3% (−10.2%, 0.8%) | 1.3% (−7.2%, 3.9%) |
| | 50% | 0.9% (−6.1%, 2.9%) | 4.5% (−0.9%, 9.9%) |
| | 85% | 1.9% (−2.0%, 4.7%) | 6.9% (2.5%, 14.8%) |
| (C) Assuming that the one-dose schedule provided 30-year protection [c] | | | |
| (C)(i) 2F1M | 25% | 1.7% (0.5%, 2.9%) | 4.7% (1.6%, 7.6%) |
| | 50% | 2.5% (0.8%, 5.6%) | 6.9% (2.5%, 13.0%) |
| | 85% | 3.2% (1.2%, 6.7%) | 8.4% (3.1%, 16.0%) |
| (C)(ii) 1F1M | 25% | 0.7% (−1.9%, 1.7%) | 3.5% (1.0%, 6.2%) |
| | 50% | 2.0% (0.5%, 4.4%) | 6.6% (2.1%, 12.4%) |
| | 85% | 2.9% (1.0%, 6.5%) | 8.1% (3.1%, 15.9%) |

Abbreviations: FOV, female-only vaccination; GNV, gender-neutral vaccination; xFyM, x-dose schedule for schoolgirls and y-dose schedule for schoolboys; 2dFOV, two-dose schedule for FOV.

Notes. [(a)] For each GNV or FOV strategy, we first calculated the relative change in the number of genital warts compared to no HPV vaccination, i.e., $\Delta GW_{\text{vax}} = 1 - GW_{\text{vax}}/GW_{\text{novax}}$, where $GW_{\text{vax}}$ refers to the estimated number of genital warts in a vaccination strategy and $GW_{\text{novax}}$ to the situation when no HPV vaccination is available. Then, we estimated the additional relative reduction of a GNV strategy compared to 2dFOV from $\Delta GW_{\text{GNV}} - \Delta GW_{\text{FOV}}$. Cases of genital warts among age cohorts involved in the vaccination program over the time horizon of 100 years were considered. [(b)] All GNV strategies were compared to the status quo 2dFOV. Vaccine uptake among girls was set at 85% in all vaccination strategies. [(c)] The two-dose schedule was assumed to provide lifelong protection. The duration of vaccine protection of the one-dose schedule was assumed at 20 years in (B) and 30 years in (C). [(d)] The cells present the medians and 90% prediction intervals (5$^{\text{th}}$ to 95$^{\text{th}}$ percentiles) for the additional relative reduction. A negative value of the relative reduction for 1F1M represents that 1F1M prevented fewer genital warts cases than 2dFOV.
